# A rapid review of the effectiveness of interventions and innovations relevant to the Welsh NHS context to support recruitment and retention of clinical staff

**DOI:** 10.1101/2022.05.11.22274903

**Authors:** Deborah Edwards, Judit Csontos, Elizabeth Gillen, Judith Carrier, Ruth Lewis, Alison Cooper, Micaela Gal, Rebecca-Jane Law, Jane Greenwell, Adrian Edwards

## Abstract

The National Health Service (NHS) is experiencing an acute workforce shortage in every discipline, at a time when waiting times are at a record high and there is a growing backlog resulting from the COVID-19 pandemic. This Rapid Review aimed to explore the effectiveness of interventions or innovations relevant to the Welsh NHS context to support recruitment and retention of clinical staff. The review is based on the findings of existing reviews supplemented by a more in-depth evaluation of included primary studies conducted in the UK or Europe. The review identifies a range of interventions that can be used for enhancing recruitment and retention in Wales, particular in rural areas, and supports multiple-component interventions. The findings highlight the importance of providing and locating undergraduate and post graduate training in rural locations. The findings also corroborate the use of bursary schemes for training, such as those already available for Nursing in Wales. Further, more robust evaluations, based on comparative studies, are required to assess the effectiveness of interventions to support recruitment and retention of clinical staff. There was limited evidence on interventions aimed at allied health professionals. Most of the primary studies included in the reviews used cohort (pre-post test) or cross-sectional designs. Most studies lacked a comparison group and did not use statistical analysis.

**TOPLINE SUMMARY:** *What is a Rapid Review?:* Our rapid reviews use a variation of the systematic review approach, abbreviating or omitting some components to generate the evidence to inform stakeholders promptly whilst maintaining attention to bias. They follow the methodological recommendations and minimum standards for conducting and reporting rapid reviews, including a structured protocol, systematic search, screening, data extraction, critical appraisal, and evidence synthesis to answer a specific question and identify key research gaps. They take 1-2 months, depending on the breadth and complexity of the research topic/ question(s), extent of the evidence base, and type of analysis required for synthesis. This report is linked to **a prior rapid evidence map** published as: What innovations (including return to practice) would help attract, recruit, or retain NHS clinical staff? A rapid evidence map, report number – REM00028 (May 2022)

*Background / Aim of Rapid Review:* The National Health Service (NHS) is experiencing an **acute workforce shortage** in every discipline, at a time when **waiting times are at a record high** and there is a growing **backlog** resulting from the **COVID-19 pandemic**. This Rapid Review aimed to explore the **effectiveness of interventions or innovations** relevant to the Welsh NHS context to **support recruitment and retention of clinical staff**. The review is based on the findings of existing reviews supplemented by a more in-depth evaluation of included primary studies conducted in the UK or Europe.

*Key Findings:* Extent of the evidence base

- 8 systematic reviews and 1 scoping review (with an evaluation component) were included. The reviews included 292 primary studies (218 unique studies), 9 of which were conducted in Europe and UK.
- The reviews focused on dentists (n=1), general practitioners (n=1), physicians (n=1); the medical workforce including undergraduates (n=1), medical undergraduates (n=1), and a variety of different health professionals (n=3) including those in training (n=1).
- Most reviews (n=8) looked for evidence of interventions within rural, remote or underserved areas.
- The interventions were mapped across categories described by the WHO (2010). Recency of the evidence base
Most of the primary studies (n=275) were conducted within the last 20 years.

*Evidence of effectiveness:* Educational interventions (8 reviews):

- **Selecting students based on rural background**: **positive association** with *recruitment* and *retention* (moderate-low quality evidence from 5 reviews).
- **Locating education institutions in rural areas** / providing training within rural oriented medical schools: **positive association** with *recruitment and retention* (low quality evidence from 3 reviews).
- Exposure to **rural health topics as part of the taught curricula** for undergraduates and postgraduates: **positive association** with *recruitment* (moderate-low quality evidence from 2 reviews).
- **Rural clinical placements**, fellowships or internships in undergraduate or post-graduate education: **mixed evidence** associated with rural intentions or actual employment (*recruitment and retention*; low quality review evidence from 7 reviews).
- **Facilitating continuing education** for rural and remote healthcare professionals: positive association with rural *recruitment and retention* (low quality evidence from 2 reviews).
- **‘Rural-based training programmes’**: **positive association** for doctors and healthcare professionals (Moderate quality evidence from 2 reviews) with rural *recruitment and retention*. Regulatory interventions requiring return to service in rural areas (6 reviews):

- Bonded schemes, **scholarships or bursaries**: **positive association** with *recruitment* but not *retention* (Low quality evidence from 2 reviews)
- **Visa Waivers**: **mixed evidence** on *recruitment and retention* (4 reviews)
- **Financial incentives**: **mixed evidence** (1 review)
- **Loan repayments**: associated with high *retention* (low quality evidence from 1 review)
- **Access to professional licences** and/or provider number for international medical graduates: associated with *low retention* (low quality evidence from 1 review)
- **Accelerated clinical training**: positive association with *retention* (low quality evidence from 1 review)
- **Enhance scope of practice**: **positive association** with *retention* (low quality evidence from 1 review)
- **Compulsory service**: **effective/positive association** with *retention* (low quality evidence from 2 reviews)
- National Health Insurance scheme: effective in terms of *recruitment and retention* (low certainty review evidence from 1 review; only one small study identified) Financial incentives without return to service requirement (3 reviews):

- **Benefits that make working in rural areas** more attractive and offset other costs/losses (e.g. higher salaries) **or in-kind benefits** (e.g. subsidised or free housing or vehicles): **inconclusive evidence** for high income countries, but positive association in middle income countries for improving *recruitment* and *retention* (low quality evidence from 3 review). A very low-quality UK study reported a **positive association**.
- **Loan re-payment programmes: positive association** with *retention* (low quality evidence from 1 review) Personal and professional support – factors that improve living and working conditions in rural areas (3 reviews):

- Positive association with *retention* (low level evidence from 3 reviews) Bundled strategies (4 reviews):

- There was consensus that multi-component interventions positively impacted on *recruitment,* and *retention* of rural workforce Policy Implications

- The review identifies a range of interventions that can be used for enhancing recruitment and retention in Wales, particular in rural areas, and supports multiple-component interventions.
- The findings highlight the importance of providing and locating undergraduate and post graduate training in rural locations.
- The findings corroborate the use of bursary schemes for training, such as those already available for Nursing in Wales.
- Further, more robust evaluations, based on comparative studies, are required to assess the effectiveness of interventions to support recruitment and retention of clinical staff. There was limited evidence on interventions aimed at allied health professionals. Strength of Evidence
Most of the primary studies included in the reviews used cohort (pre-post test) or cross-sectional designs. Most studies lacked a comparison group and did not use statistical analysis.

## 1. BACKGROUND

This Rapid Review is being conducted as part of the Wales COVID-19 Evidence Centre Work Programme. The above question was suggested by the Royal College of Surgeons, Edinburgh.

### 1.1 Purpose of this review

National Health Service (NHS) waiting times have significantly increased over the past couple of years, particularly since the emergence of COVID-19, as elective and non-emergency treatments have been suspended or delayed to focus on the pandemic response. In Wales, 240,306 people were waiting more than 36 weeks for treatment from referral in September 2021, thirteen times more than in September 2019 (Welsh Government 2021). However, clearing the backlog is difficult, as there is NHS workforce shortage in every speciality, with 93,000 job vacancies UK-wide (Health and Social Care Committee 2021).

For years, there has been an observable tendency that lower number of healthcare professionals enter the NHS than the number of qualified workers leaving, contributing to workforce shortages (Health Committee 2018). Several factors are responsible for the NHS staff retention issues, including excessive workload, antisocial working patterns, physical and emotional strain, stress, burnout, stigma, and negative portrayal of healthcare work in the media (Darbyshire et al. 2021, Mitchell et al. 2021, Royal College of Anaesthetists 2021). While these issues predate the pandemic, they have been exacerbated by the increased pressures rising COVID-19 infections and hospitalisations meant (Falatah 2021), overstretching an already limited NHS staff. Factors contributing to retention and recruitment issues need to be addressed, and numerous strategies will be required to fill the workforce gap (British Medical Association 2021).

This rapid review is an extension of a previously completed rapid evidence map published as: What innovations (including return to practice) would help attract, recruit, or retain NHS clinical staff? A rapid evidence map, report number – REM00028 (April 2022). This report is available from the WCEC library: https://healthandcareresearchwales.org/wales-covid-19-evidence-centre-report-library. The evidence map was used to identify the extent and nature of the available evidence and encompassed a broad range of innovations and factors that would help attract, recruit, or retain NHS clinical staff. In preparing the rapid evidence map we found a number of systematic reviews based on international literature that focussed on innovations that might help recruit and retain the healthcare workforce. The findings of the evidence map were presented to the stakeholders, and a decision made that the rapid review should focus on evidence transferable to the Welsh context. The evidence map identified that there was sufficient evidence to undertake a rapid review of reviews in this area.

This rapid review aimed to explore the effectiveness of interventions or innovations relevant to the Welsh NHS context to support recruitment and retention of clinical staff.

## 2. RESULTS

### 2.1 Overview of the evidence base

Of the 5056 citations retrieved from our searches, ten met our eligibility criteria, which included eight systematic reviews (across nine publications) and one scoping review with an evaluative component. The evidence review conducted by the World Health Organisation (WHO), which represents an update review, was reported across two publications (WHO 2020, WHO 2021). This systematic review covered the time period 2010 to 2019 (WHO 2020) and represents an update of an earlier review published in 2010, which covered the time period 1995 to 2009 (WHO 2010). The details of the methods included studies, and Grading of Recommendations Assessment, Development and Evaluation (GRADE) evidence profiles for the update review can be found in the 2020 publication (WHO 2020). However, this document only provides a basic summary of the evidence and data were also extracted from the 2021 WHO guideline document (WHO 2021). The authors state that this guideline is based on the combined primary source evidence from both the 2010 and 2020 WHO reviews.

For full details of the nine included systematic reviews can be found in Table 1. A total of 292 primary studies were cited by the included systematic reviews, 74 of which were duplicated across the systematic reviews (see Section 5.6). This represents a slight overlap, with the systematic reviews mostly considering different primary studies (Pieper et al. 2014). The recency of the evidence base is as follows: 1970s (n=3); 1980s (n=1); 1990s (n=13), 2000s (n=275).

### 2.2 Description of the included reviews

The included systematic reviews focused on dentists (Suphanchaimat et al 2016), general practitioners (GPs) (Verma et al. 2016), physicians (Kumar and Clancy 2020); the medical workforce including undergraduates (Noya et al. 2021), medical undergraduates (Johnson et al. 2018), and a variety of different healthcare professionals (Grobler et al. 2015, Russell et al. 2021, WHO 2020, 2021) including those in training (MacQueen et al. 2018).

One systematic review evaluated strategies used within primary care in high income countries (Verma et al. 2016). A further eight systematic reviews looked for evidence of interventions within rural, remote, or underserved areas. However, there was no consistency in the definition of such areas with authors either using national or international classification systems (n=2) (Russell et al. 2021, WHO 2020, 2021), as defined within the primary studies (n=5) (Grobler et al. 2015, Kumar and Clancy 2020, Macqueen et al. 2018, Noya et al. 2021, Suphanchaimat et al. 2016) or did not provide any further details (n=1) (Johnson et al. 2018). As there was no consensus in the definition of rural in the included systematic reviews, in this rapid review of reviews the ’term’ rural is used according to the individual review author’s definition, thus encompass rural, remote, and underserved areas.

Systematic reviews sought to include studies either from countries of all income levels (n=5: Grobler et al. 2015, Johnson et al. 2018, Kumar and Clancy 2020, Suphanchaimat et al. 2016, WHO 2020, 2021); from high income countries (HICs) (n=3: MacQueen et al. 2018 (USA only), Russell et al. 2021, Verma et al. 2016) and Noya et al. (2021) looked at similarities and differences between HICs and low- and middle-income countries (LMICs).

Across all the included systematic and scoping reviews, no study reported using a randomised controlled trial to assess a strategy. Twelve of the included studies across five of the systematic reviews (Johnston et al. 2018, MacQueen et al. 2018, Russell et al. 2021, Verma et al. 2016, WHO 2020, 2021) and the scoping review (Noya et al. 2021) used quasi experimental (n=4) or pre-post test (n=8) designs. The majority of the studies within the systematic reviews were cohort (pre-post test, prospective or retrospective) studies or cross-sectional designs.

**<u>Recruitment</u> was measured in the included systematic reviews as the proportion of healthcare professionals who initially choose to work in a designated clinical area** as a consequence of being exposed to the intervention (n=6: Grobler et al. 2015, MacQueen et al. 2018, Noya et al. 2021, Suphanchaimat et al. 2016; Verma et al. 2016, WHO 2020, 2021) **or who have future intentions of working in a designated clinical area** (n=3: Johnson et al. 2018, Suphanchaimat et al. 2016, Verma et al. 2016).

**<u>Retention</u> was measured as either the proportion of healthcare professionals who continue to work in a designated clinical area** as a consequence of being exposed to the intervention (n=8: Grobler et al. 2015, Kumar and Clancy 2020, MacQueen et al. 2018, Noya et al. 2021, Russell et al. 2021, Suphanchaimat et al. 2016, Verma et al. 2016, WHO 2020, 2021) or health professional **preferences** (n=1: Russell et al. 2021) or **intentions to remain in their current job** (n=4: Noya et al. 2021, Russell et al. 2021; Suphanchaimat et al. 2016; Verma et al. 2016).

The interventions to recruit and retain clinical staff were mapped across the categories as described by the WHO (2010) which are:

- Educational interventions (Johnson et al. 2018, Kumar and Clancy 2020, MacQueen et al. 2018, Noya et al. 2021, Russell et al. 2021, Suphanchaimat et al. 2016, Verma et al. 2016, WHO 2020, 2021).
- Regulatory interventions (Grobler et al. 2015, Kumar and Clancy 2020, Noya et al. 2021, Russell et al. 2021, Verma et al 2016, WHO 2020, 2021).
- Financial incentives (Kumar and Clancy 2020, Russell et al. 2021, WHO 2020, 2021).
- Personal and professional support (Kumar and Clancy 2020, Russell et al. 2021, WHO 2020, 2021).
- Bundled strategies (Kumar and Clancy 2020, Noya et al. 2021, Verma et al. 2016, WHO 2020, 2021).

Other innovations that were evaluated across the evidence base but did not map to the WHO categories included international recruitment, marketing, retainer schemes, re-entry schemes, specialised recruiters or case managers and delayed partnerships (Verma et al. 2016) and those related to health systems (Russell et al. 2021).

### 2.3 Description of primary studies conducted within the UK and Europe

The included reviews were searched for primary research studies conducted in Europe and UK that could be transferable to the Welsh context. More details on how primary studies were selected can be read in the methods (Section 5.6). Searches identified nine primary research studies conducted within Europe and the UK. Professions of interest were doctors including GPs (n=5) (Chevillard et al. 2019, Flum et al. 2016, Gaski and Abelsen 2017, MacVicar et al 2016, Straume and Shaw 2010), nurses (n=2) (Nilsen et al. 2012, Norbye and Skaalvik 2013), Allied health professionals (AHPs) (n=1) (Solowiej et al. 2010) and mixed group of healthcare professionals (n=1) (Carson et al. 2015). The countries where the research was conducted were Norway (n=4) (Gaski and Abelsen 2017, Nilsen et al. 2012, Norbey and Skaalvik 2013, Straume and Shaw 2010), UK (n=2) (MacVicar et al. 2016, Solowiej et al. 2010), France (n=1) (Chevillard et al. 2019), and Germany (n=1) (Flum et al. 2016). One further study investigated strategies across six European countries (Ireland, Scotland, Iceland, Greenland, Norway, and Sweden). All primary research studies were conducted within rural, remote, or underserved areas (Carson et al. 2015).

Study designs included descriptive surveys (n=4) (Carson et al. 2015, Flum et al. 2016, MacVicar et al. 2016, Norbye and Skaalvik 2013), quantitative methods using retrospective datasets (n=2) (Chevillard et al. 2019, Straume and Shaw 2010), and a cohort study (n=1) (Gaski and Abelsen 2017). Two further studies were part of a wider mixed methods approach and as a quantitative element used descriptive surveys (n=1) (Solowiej et al. 2010) and cohort study design (n=1) (Nilsen et al. 2012).

### 2.4 Quality of the included reviews

Included systematic reviews and the scoping review with an evaluative component were critically appraised with the JBI checklist for systematic reviews and research syntheses (Aromataris et al. 2015) (further information on the JBI checklist and the quality appraisal is in the additional material which is available on request from the WCEC). None of the systematic reviews met all 11 quality criteria of the JBI checklist. Out of the eight systematic reviews, only one met 10 quality criteria (Grobler et al. 2015), indicating an overall good quality due to the absence of reporting of publication bias assessment.

Four studies met nine quality criteria out of 11 (Johnson et al. 2018, Verma et al. 2016, Russell et al. 2021, WHO 2020, 2021) with common methodological issue being a lack of publication bias assessment. Other quality issues included vague description of methods, such as the number of reviewers conducting quality assessment (Johnson et al 2018) or data synthesis approach (Verma et al. 2016). The absence of recommendations for either policy and practice or future research were also identified as problematic for two systematic reviews (Russell et al. 2021, WHO 2020, 2021).

One systematic review met eight out of 11 quality criteria (MacQueen et al. 2018), with issues including a lack of clarity around how quality appraisal and publication bias assessment was conducted, and the absence of recommendations for future research. One systematic review met seven quality criteria (Kumar and Clancy 2020) due to unclear description of their search strategy, insufficient number of resources and databases searched, and lack of clarity on publication bias assessment. One systematic review met six quality criteria out of 11 due to insufficient risk of bias assessment, lack of clarity in data extraction methods, and the absence of clear recommendations for policy and practice and for future research (Suphanchaimat et al. 2016). The scoping review with evaluative component met six out of a possible seven quality criteria (Noya et al. 2021) due to a lack of clarity in how data extraction was conducted.

### 2.5 Effectiveness of educational interventions

Seven systematic reviews and one scoping review with an evaluative component investigated the effectiveness of a number of different educational interventions.

- Selecting health professional students based on rural background (Russell et al. 2021, Suphanchaimat et al. 2016, Noya et al. 2021, Verma et al. 2016, WHO 2020, 2021).
- Locating education institutions in rural areas / providing training within rural oriented medical schools (Johnson et al. 2018, Noya et al. 2021, WHO 2020, 2021).
- Exposure to rural health topics as part of the taught curricula for undergraduates and postgraduates (Johnson et al. 2018, WHO 2020, 2021).
- Rural clinical placements, fellowships, or internships in undergraduate or post- graduate education (Johnson et al. 2018, Kumar and Clancy 2020, Russell et al. 2021, Suphanchaimat et al. 2016, Noya et al. 2021, Verma et al. 2016, WHO 2020, 2021).
- Facilitating continuing education for rural and remote healthcare professionals (Russell et al. 2021, WHO 2020, 2021).
- ‘Rural-based training programmes’ which is an overarching term that covers interventions, such as rural residency training, rural campus, association with rural medical schools, and rural summer externships among others (MacQueen et al. 2018, Kumar and Clancy 2020).

### 2.6 Evidence from reviews for doctors (including general practitioners)

One systematic review explored the evidence base for rural educational programmes within **medical education**, specifically rural clinical **placement programmes** with and without rural health educational curriculum and **rural clinical school programmes**. **Mixed evidence** was reported for the effectiveness of such interventions on **rural intentions or actual rural employment** with 55% of studies providing strong evidence of an effect (Johnson et al. 2018). Kumar and Clancy (2020) summarised the recent literature that analysed strategies implemented to increase retention for physicians and reported that effective strategies were rural-based training programmes for students training in HICs and rural placements for students training in MICs. Verma et al. (2016) reported weak evidence for an association between the improvement in the **recruitment** of **primary care doctors** and postgraduate placements in underserved areas, undergraduate rural placements and recruiting students to medical school from rural areas. Noya et al. (2021) found that educational strategies which included rural background student selection, rural exposure during medical school, and rural oriented medical school positively impacted recruitment and retention of the rural medical workforce.

### 2.7 Evidence from reviews for dentists

Suphanchaimat et al. (2016) included a meta-analysis and found that **dental students/graduates** with rural experience (defined as increased exposure to rural areas during training or recruiting students from a rural background) were four times as likely to have intentions to practice in rural areas than those without rural experience (OR 4.06 (95% CI, 2.55–6.45). Subgroup analysis showed that the teaching programmes that included greater exposure to rural areas tended to have a marginally greater impact on intention to practice in rural areas compared to interventions that aimed to recruit more students with a rural background (OR 4.32: 95% CI, 1.93-9.66 versus OR 4.20; 95% CI, 2.22–7.96), but both strategies still had a statistically significant impact.

### 2.8 Evidence from reviews for mixed groups of healthcare professionals

The success rate of US rural training programmes was found to range from 30 to 65% in the systematic review conducted by MacQueen et al. (2018). This was based on moderate quality evidence and on average it was reported that just one in every two trainees are likely to enter rural care after having undertaken a rural training programme. Russell et al. (2021) demonstrated that preferential selection of students who grew up in a rural area, undertaking substantial lengths of rural training during basic university training or during post-graduate training and supporting existing rural health professionals to extend their skills or upgrade their qualifications was associated with increased rural retention. This was based on low quality evidence. The systematic review and subsequent guidelines published by the WHO (WHO 2020, 2021) found moderate quality evidence of the effectiveness of enrolling students with a rural background in health worker education programmes. Admitting students with a rural background has a positive effect on the availability of rural health workers. Additionally, low quality evidence was reported for locating education facilities closer to rural areas, exposure to rural health topics as part of the taught curricula for undergraduate and postgraduate, rural clinical placements, aligning health worker education with rural health needs and facilitating continuing education for rural and remote healthcare professionals.

### 2.9 Evidence from primary research conducted within the UK and Europe

From the included systematic and scoping reviews, seven primary studies investigated educational interventions in Europe and UK (Carson et al. 2015, Flum et al. 2016, Gaski and Abelsen 2017, MacVicar et al. 2016, Nilsen et al. 2012, Norbye and Skaalvik 2013, Straume and Shaw 2010). These were conducted in Norway (n=4), Scotland (n=1), Germany (n=1), and across seven European countries (Ireland, Scotland, Iceland, Greenland, Norway and Sweden) (n=1) and study designs included four descriptive surveys (n=4), a quantitative study using retrospective datasets (n=1), a cohort study (n=1), and a cohort study within mixed methods design (n=1). The following interventions were investigated:

- Rural clinical placements, fellowships or internships in undergraduate or post- graduate education (Gaski and Abelsen 2017, MacVicar et al 2016, Straume and Shaw 2010).
- Exposure to rural health topics as part of the taught curricula for undergraduates and postgraduates (Flum et al 2016).
- Locating education institutions in rural areas (Nielsen et al. 2012, Norbye and Skaalvik 2013).
- Selecting health professional students based on rural background (Carson et al. 2015).

Flum et al. (2016) investigated whether a “rural day” was an effective intervention for GP workforce shortages in rural communities in Germany. The ‘rural day’ programme was a day trip to rural communities with presentations by political stakeholders about the programme, evidence, and strategies; information on the rural region; informal discussions between GP trainees and political stakeholders; visits to primary care service and local points of interest. The results showed that the rural day had no significant influence on intention to work in rural practice but that there was an increase in positive attitudes towards rural areas in general.

The impact of primary care internships on recruitment and retention to rural and remote areas of Norway was reported by Straume and Shaw (2010). The results indicated that a primary care internship might have a positive impact on vacancy rate, although no statistical analysis was presented to confirm the findings. A further study investigated the choice of workplace following the same internship programme after the introduction of an early sign-up model which gave students in their tenth term the opportunity to sign up for the internship placement (Gaski and Abelsen 2017). It was expected that interns who chose to work in rural areas would have a more positive attitude towards working within the rural study area following the internship than those who were assigned a regular internship placement. The results showed that physicians with an early sign-up internship who remained working in the study area was nearly double compared to physicians with a regular internship. However, this only appeared to be the case for the most densely populated areas and not the regions with smaller or more scattered populations.

Two further studies from Norway investigated whether decentralized nursing education that provides off-campus training in rural areas contributed to recruitment and retention (Nilsen et al. 2012, Norbye and Skaalvik 2013). Norbye and Skaalvik (2013) presented the results of a survey and showed that the off-campus programme had been successful in recruiting and retaining nurses to rural areas. Nilsen et al. (2012) conducted a cohort study, which reported that the off-campus classes resulted in a considerably higher retention rate (92.5%) compared to the traditional campus classes retention rate (70%). However, no statistical analysis was presented to confirm the findings.

MacVicar et al. (2016) explored the impact of a GP rural fellowship programme in which GPs in Scotland were offered a further year of training in and exposure to rural medicine. The results of a survey found that just under three quarters of the graduates who had completed the fellowship programme were retained in rural roles.

One further descriptive study (Carson et al. 2015) looked at the impact of the ‘rural pipeline’ on retention of doctors, nurses and AHPs across seven European countries (Ireland, Scotland, Iceland, Greenland, Norway and Sweden). This study was included in the WHO review even though it did not evaluate an intervention rather it explored the concept of “rural pipeline”. It has however, been included in here for completeness. The rural pipeline includes ‘rural origin’ (those who grew up in rural areas to enter the health professions) and training in rural locations, and other forms of exposure, such as visits to rural communities (rural exposure). The research concluded that overall, the rural pipeline (both rural origin and rural exposure) does impact on retention. However, when relationships between rural pipeline and retention were explored by country, positive association for healthcare professionals in Scotland, UK were no longer present.

### 2.10 Bottom line summary for educational interventions

This section summarised evidence from seven systematic reviews and one scoping review with an evaluative component, three for doctors including GPs, one for medical workers including undergraduates, one for dentists and three for mixed groups of healthcare professionals.

- **Mixed evidence** was found for **trainee doctors (including GPs)** of an association between **rural clinical placements** in both undergraduate and postgraduate education, rural health educational curriculum, rural clinical school training programmes and **rural intentions or actual rural employment.**
- **Moderate quality evidence** from one scoping review with an evaluation component for the medical workforce demonstrated a **positive association** between selecting students based on rural background, encouraging undergraduate and postgraduate training exposure to rural areas as part of the curricula and **rural recruitment and retention.**
- **Moderate quality evidence** suggested that one in every two trainee healthcare professionals are likely to **enter rural care** after having undertaken a rural training programme.
- **Low quality evidence** demonstrated a **positive association** between selecting **healthcare students (including dentists)** based on rural background and recruitment in rural areas. One further systematic review also found low quality evidence of a positive association between exposure to rural health topics as part of the taught curricula for undergraduates and postgraduates and **recruitment in rural areas.**
- **Moderate quality evidence** from one systematic review demonstrated a **positive association** between selecting **healthcare** students based on rural background and **recruitment and retention in rural areas.**
- **Low quality evidence** from one systematic review across healthcare professionals demonstrated a positive association between locating education facilities closer to rural areas, bringing students in health worker education programmes to rural and remote communities, aligning health worker education with rural health needs, facilitating continuing education for rural and remote healthcare professionals and **recruitment to rural areas.** In addition, from the same review **low quality evidence** suggested a **positive association** between facilitating continuing education for rural and remote healthcare professionals **and retention.**

Evidence was also summarised for seven of the primary studies that had been conducted within the UK and Europe.

- **Low and very low quality** evidence from three primary studies suggested a **positive association** between internships and fellowships for GPs and GP trainees and **recruitment and retention** in rural areas.
- **Low quality** evidence from one primary study showed that rural exposure in the form of a ‘rural day’ had **no impact on intentions** to work in rural areas, although GP trainees’ attitude towards these locations improved.
- **Low and moderate evidence** from two primary studies suggested a **positive association** between off-campus undergraduate education and **nurse retention**.
- **Low quality evidence** from one primary study conducted across several European countries with **healthcare professionals** indicated that overall ‘rural pipeline’ (rural origin or rural exposure) can have a **positive impact on retention**, although results specific to the UK showed no positive influence on retaining healthcare professionals.

### 2.11 Effectiveness of regulatory interventions

Five systematic reviews and one scoping review with an evaluative component investigated the effectiveness of a number of different regulatory interventions requiring an obligatory return of service (RoS) in a rural area

- Bonded schemes, scholarships, or bursaries - subsidies to attend education and development events over the course of the program, relocation allowances to assist with moving for their rural service in return for a RoS contract to complete a predefined number of postgraduate years in a rural hospital (Noya et al. 2021, WHO 2020, 2021).
- Visa waivers - obligatory service schemes linked to work in rural areas in return for concessions on international medical graduates (IMGs) visa requirements (Kumar and Clancy 2020, Noya et al. 2021, Russell et al. 2021, Verma et al. 2016).
- Loan repayments (Russell et al 2021).
- Access to professional licences and/or provider number for IMGs (Russell et al 2021).
- Enhanced scope of practice in rural areas (WHO 2020, 2021).
- Introduction of different types of health workers with the appropriate training such as accelerated medically trained clinicians (WHO 2020, 2021).
- Compulsory service – mandatory deployment of health workers (Kumar and Clancy 2020, WHO 2020, 2021).
- National Health Insurance scheme (Grobler et al 2015).
- Other financial incentives with RoS (Verma et al. 2016)

### 2.12 Evidence from reviews for doctors (including general practitioners)

Noya et al. (2021) reported evidence that bonded scholarships and visa waiver programmes for IMGs positively impacted recruitment in rural areas for the medical workforce. However, Kumar and Clancy (2020) reported that for IMGs in HICs visa waiver programmes were identified as a non-effective retention strategy. The same review reported that compulsory rural service programmes for physicians were effective as a retention strategy in HICs but that the evidence was inconclusive within MICs (Kumar and Clancy 2020). One further systematic review reported that evidence base for financial incentives with RoS was mixed (Verma et al. 2016). The quality of studies was not sufficient to make any firm conclusions regarding the effectiveness of visa waiver programmes for IMGs in rural areas, although all studies did report success in recruiting international doctors, but retention rates varied (Verma et al. 2016).

### 2.13 Evidence from reviews for mixed groups of healthcare professionals

Russell et al. (2021) found that evidence regarding interventions which required service in rural areas (for a varying length of time) in return for a benefit (exchange for visa waivers, access to professional licenses or provider numbers) were associated with comparatively low rural retention, especially once the RoS period was complete. However, rural retention was higher if RoS was in exchange for loan repayments.

Scope of practice can be defined as services a healthcare professional based on their level of experience and competence is allowed to provide within the limits of their professional and regulatory standards, registration, qualification, and the approval of their organisation. Due to limited availability of different health professions and specialities in rural and remote areas, existing healthcare professionals are often required to provide services beyond the scope of their practice, thus risking quality of care (WHO 2020, 2021). An enhanced scope of practice is defined as “the development or acquisition of skills or expertise beyond the currently recognized scope of practice” (WHO 2021, p.10). The WHO (2020, 2021) reported that the introduction of an enhanced scope of practice increases job satisfaction which positively influences retention and improves access to health care for rural communities with shortages of high-level health workers or specialists (GRADE – low).

Another successful solution to workforce shortages reported in the WHO review (2020, 2021) is the recruitment of health workers in rural areas, that are faster to train and more readily deployed and retained in rural areas (for example, accelerated medically trained clinicians) (GRADE – low). There was also low certainty of evidence regarding the impact of compulsory service, and the evidence mainly focused on doctors and dentists, and the impact is therefore limited. As an alternative to compulsory service, tertiary education providers in HICs offer health professions scholarships, bursaries, stipends or other forms of subsidies to cover the costs of their education and training in return for an agreement to work in a rural or remote location for a certain period after qualification. The WHO review (2020, 2021) demonstrated positive influences on the increase in the availability of service-obligated health workers through high rates of completion of the service agreements and varied retention rates (GRADE – low).

The systematic review by Grobler et al. (2015) only retrieved one study and this evaluated the implementation of National Health Insurance (NHI) scheme on the distribution of health professionals in Taiwan. Prior to the implementation of the NHI, people living in urban areas were more likely to be able to afford higher medical costs. Therefore, it was more financially rewarding for health providers to work in urban areas compared to the poorer rural areas. However, due to very low certainty of the evidence it was not possible to be certain about the effect of NHI schemes on the distribution of health professionals.

### 2.14 Evidence from primary research conducted within UK and Europe

We did not find any evidence from primary studies conducted within the UK and Europe that explored regulatory interventions within the included reviews.

### 2.15 Bottom line summary for regulatory strategies

This section summarised evidence from five systematic reviews and one scoping review with an evaluative component, two for doctors including GPs, one for medical workers including undergraduates, and three for mixed groups of healthcare professionals.

- **Low quality evidence** from one systematic review reported an association between RoS interventions in rural areas (visa waivers, access to professional licenses or provider numbers) for IMGs across a range of healthcare professional groups and low rural retention.
- **Low quality evidence** was found for an association between RoS interventions in rural areas (loan repayments) across a range of healthcare professional groups and high rural retention. There was **mixed evidence** for the effectiveness of **visa waiver programmes** on the **recruitment and retention o**f **doctors (including GPs) in rural areas**.
- There was mixed evidence for the effectiveness of **financial incentives with RoS** on the **recruitment and retention o**f **doctors (including GPs) in rural areas**. Evidence from one scoping review with an evaluative component reported that **bonded scholarships positively impacted** recruitment of the medical workforce in rural areas.
- One systematic review demonstrated that compulsory rural service programmes for **physicians** were effective for improving retention **in HICs.**
- **Low quality evidence** from one systematic review reported a positive association between **an enhanced scope of practice**, accelerated medical clinicians training, compulsory service programmes and increase in the availability of service-obligated health workers (scholarships, bursaries, stipends or other forms of subsidies), and the **retention** of healthcare professionals.
- One study within one systematic review found low certainty evidence for the effect of NHI schemes on the distribution of healthcare professionals.

### 2.16 Effectiveness of financial incentives

Three systematic reviews reported on the effectiveness of financial incentives without RoS (Russell et al. 2021, Kumar and Clancy 2020, WHO 2020, 2021). Financial incentives include:

- Monetary benefits that offset other costs and losses of working rurally, such as higher salaries; and in-kind benefits such as, subsidised school fees for children, subsidised or free housing or vehicles, smartphones, post graduate training opportunities (Russell et al. 2021, WHO 2020, 2021)
- Loan payment programmes without RoS (Kumar and Clancy 2020).

### 2.17 Evidence from reviews for doctors (including general practitioners)

Kumar and Clancy (2020) reported that for physicians in MICs financial incentives were found to be effective as retention strategies. However, in HICs the effectiveness of financial incentives was inconclusive, although it was suggested that direct financial incentives without RoS are most effective, particularly loan repayment programs (in countries with high tuition fees for medical education).

### 2.18 Evidence from reviews for mixed groups of healthcare professionals

Russell et al. (2021) reported that the evidence about the impact of financial incentives (with no RoS requirement) for healthcare professionals was limited because of the small number of studies and insufficient methods used to capture and report actual retention numbers. The systematic review conducted by WHO (2020, 2021) reported that findings from some observational studies suggest that salaries and allowances are positively linked to the recruitment and retention of health workers in rural areas in both the short and medium term (GRADE – low). Other non-monetary incentives have been shown to positively influence job satisfaction.

### 2.19 Evidence from primary studies conducted within the UK and Europe

One descriptive study assessed the impact of an AHP support and development scheme funding package of £3000 on the recruitment and retention of AHPs working in rural areas of NHS Scotland. The managers of the scheme reported that the majority (89%) of hard to fill vacancies had been filled and that new team members had stayed in the post for an average of one and a half years (Solowiej et al. 2010).

### 2.20 Bottom line summary for financial incentives

This section summarised evidence from three systematic reviews, one for doctors and two for mixed groups of healthcare professionals.

- There was **low quality mixed evidence** for the effectiveness of **financial incentives** without RoS for improving recruitment of doctors and other healthcare professionals across HICs. However, one systematic review reported that financial incentives were effective retention strategies for physicians in MICs
- In rural areas, loan repayment programs (in countries with high cost of medical education), salaries and allowances are **positively linked to** the recruitment and retention of healthcare professionals.

Evidence was also summarised for one of the primary studies that had been conducted within the UK and Europe.

- **Very low quality evidence** suggested that recruitment and retention **had improved** after the introduction of an AHP development and support scheme.

### 2.21 Effectiveness of personal and professional support

Four systematic reviews evaluated the effectiveness of personal and professional support (Kumar and Clancy 2020, Russell et al 2021, Verma et al. 2016, WHO 2020, 2021). Personal and professional support has been described as relating to factors that improve living and working conditions in rural areas, including community support and family integration into the community such as good infrastructure to improve living conditions (such as accommodation, running water, electricity, roads and internet access), opportunities for social interaction, schooling for children, employment for spouses, opportunities to advance careers and to communicate and consult with peers through networks, telehealth or other approaches (Kumar and Clancy 2020, WHO 2020, 2021).

### 2.22 Evidence from reviews for doctors (including general practitioners)

The review by Kumar and Clancy (2020) reported the effectiveness of personal and professional support on retention in rural areas in both HMICs and MICS. Sustainable workplace organisation and infrastructure, and social support were effective strategies to retain doctors in HICs, while workplace infrastructure was effective in MICs. Verma et al. 2016 reported that the quality of the studies was not sufficient to draw conclusions about well-being or peer support or mixed approaches.

### 2.23 Evidence from reviews for mixed groups of healthcare professionals

Russell et al. (2021) provides narrative evidence that a cognitive behavioural coaching programme and an enhanced professional network were effective strategies for retaining healthcare professionals working in rural areas but noted that studies were very low quality due to a lack of a comparison groups.

The WHO (2020, 2021) review summarised findings from observational studies that evaluated the effectiveness of a number of personal and professional support interventions that had a positive effect on retention for healthcare professionals working in rural areas, all of which were rated low using the GRADE approach.

- *Investing in rural and remote infrastructure and services to ensure decent living conditions*
- *Safe and secure working environment for health workers in rural and remote areas*
- *Decent work that respects healthcare workers’ rights, improving working conditions*
- *Health workforce support networks (availability of telehealth, mobile health and electronic health for health workers in remote and rural areas)*
- *Career development and advancement programmes, and career pathways for health workers in rural and remote areas*
- *Development of networks, associations, and journals for health workers in remote and rural areas*
- *Social recognition measures for health workers in remote and rural areas*

### 2.24 Evidence from primary studies conducted within the UK and Europe

One quantitative study which collected data using retrospective datasets explored the effectiveness of personal and professional support within the included reviews from the UK and Europe (Chevillard et al. 2019). This was conducted in France and investigated the implementation of an organisational multidimensional strategy in which multi-professional group practices (PCTs) in the form of primary care teams (GPs working with other professionals such as midwives, dentists, paramedics, nurses, or administrative staff) was implemented. The findings suggest that **PCTs help to attract and retain GPs.** The authors concluded that this was probably through the improvement of their working conditions.

### 2.25 Bottom line summary for personal and professional support

This section summarised evidence from four systematic reviews, two for doctors including GPs and two for mixed groups of healthcare professionals.

- Evidence from one systematic review indicated that sustainable workplace organisation, improved infrastructure and social support were effective in **retaining doctors** in rural areas of HICs. In rural areas of MICs, improved workplace infrastructure can have a positive impact on retention of doctors.
- **Very low quality evidence** from one systematic review reported that behavioural coaching programmes and enhanced professional networks are effective in **retaining healthcare professionals** in rural areas.
- **Low quality evidence** from one systematic review highlighted several interventions (including investing in rural infrastructure, improved working conditions and environment, workforce support network, career development opportunities, facilitating knowledge exchange via networks and journals, social recognition measures) are effective personal and professional interventions that have a positive impact on **retention of healthcare professionals** in rural areas.
- Due to the poor quality of the studies investigating well-being or peer support no conclusions could be drawn regarding effectiveness.

Evidence was also summarised for one of the primary studies that had been conducted within the UK and Europe.

- Moderate quality evidence implied that PCTs can help **attract and retain GPs** in rural areas.

### 2.26 Bundled strategies

Three systematic reviews and one scoping review with an evaluation component investigated the effectiveness of bundled (multicomponent) interventions (Kumar and Clancy 2020, Noya et al. 2021, Verma et al. 2016, WHO 2020, 2021). The WHO (2010) review describes bundled interventions as group of evidence-based interventions put together into a package that when implemented have the potential to produce better outcomes than delivered separately.

### 2.27 Evidence from reviews for doctors (including general practitioners)

One systematic review was not able to reach any conclusions about the effectiveness of bundled interventions conducted with GPs due to the poor quality of the included studies (Verma et al. 2016). Kumar and Clancy (2020) concluded that implemented strategies for physicians in the rural context must be multifactorial or bundled as well as relevant to the local context. This was based on a narrative summary across one study and two narrative reviews. Noya et al. (2021) found that bundled strategies positively impacted on recruitment, and retention of the rural medical workforce.

### 2.28 Evidence from reviews for mixed groups of healthcare professionals

The WHO (2020, 2021) found increasing evidence that bundling interventions that are relevant to the given rural context can have a synergistic effect on recruitment and retention of healthcare professionals.

### 2.29 Evidence from primary studies conducted within the UK and Europe

We did not find any evidence from primary studies conducted within the UK and Europe that explored bundled interventions within the included reviews.

### 2.30 Bottom line summary for bundled strategies

This section summarised evidence from three systematic reviews and one scoping review with an evaluation component. Three reviews investigated interventions for doctors including GPs and one for mixed groups of healthcare professionals.

- While the evidence was mixed due to the different components across the bundled interventions, there was a consensus among the included reviews that such interventions positively impacted on recruitment and retention of all healthcare professional groups.

### 2.31 Other strategies

Two systematic reviews (Russell et al. 2021, Verma et al. 2016) investigated the effectiveness of a number of interventions that did not fall within the classification system (WHO 2010). These included international recruitment (not involving visa waivers), marketing, retainer schemes, re-entry schemes, specialised recruiters or case managers and delayed partnerships (Verma et al. 2016) and those related to health systems (Russell et al. 2021).

However, Verma et al. (2016) reported that due to the poor quality of the studies investigating these interventions they were not able to reach any conclusions about retainer schemes, re-entry schemes, international recruitment, specialised recruiters, support for professional development or research or, delayed partnerships. The review conducted by Russell et al. (2021) reported just one study that focused on health system related interventions. This study was conducted in the USA where the expansion of Medicaid and Medicare in rural areas was associated with physicians giving up their rural practices and becoming clinically inactive.

## 3. DISCUSSION

### 3.1 Summary of the findings

This rapid review of reviews aimed to determine the effectiveness of interventions/innovations relevant to the Welsh NHS context to support recruitment and retention of clinical staff. Although the evidence base is weak this review found **evidence to support a wide range of educational, regulatory, personal and professional support and bundled interventions**.

The importance of rural selection to and rural exposure within undergraduate and postgraduate healthcare education has been well established from reviews that have focused on factors that impact on rural recruitment and retention (see the Rapid Evidence Map (REM), which can be downloaded from the WCEC Library. This rapid review identified that the **most common strategies reported across the evidence base were educational interventions** which demonstrated positive associations between recruitment and retention of all health professional groups and **selecting healthcare students based on rural background, locating education institutions or providing training in rural areas, exposing students to rural health topics as part of the taught curricula and facilitating continuing education for rural and remote healthcare professionals**. The updated WHO guidelines on health workers development, attraction, recruitment and retention in rural and remote areas strongly recommends “using targeted admission policies to enrol students with a rural background in health worker education programmes” (WHO 2021, pg. xiv). In addition, the review and meta-analysis by Suphanchaimat et al. (2016) for dental students revealed that students with rural exposure tended to have a fourfold-higher chance of proceeding to or intending to serve rural populations than those without exposure to rural areas. However, the effectiveness of rural clinical placements, fellowships or internships, was mixed with some studies showing a positive impact on levels of recruitment and retention and others having no impact.

There were a wide range of effective **regulatory interventions requiring RoS in rural areas** in exchange for **bonded schemes, scholarships, bursaries, visa waivers for IMGs, loan repayments, professional licences, provider numbers and/or compulsory service. However, for GP trainees there was limited mixed evidence for visa waivers**. There was also limited evidence of the impact of financial incentives with and without a RoS component across most healthcare professional groups. Russell et al. (2021) suggests that as well as the limited numbers of studies in this area, those that are conducted fail to quantify the actual retention behaviour of health professionals. In general, financial incentives were beneficial in improving recruitment and retention in rural areas in MICs and loan payment programmes without RoS in countries where the costs of healthcare education are high.

The wider literature suggests that personal and professional support may influence professionals’ choice to work in rural, remote and underserved areas (see the Rapid Evidence Map (REM), which can be downloaded from the WCEC <u>Library</u>). This rapid review of effectiveness found **positive associations between all personal and professional support interventions ranging from infrastructure support to community support and family integration** and improved recruitment and retention.

Although there was mixed evidence presented for the benefit of bundles (multifactorial interventions), there was a consensus that such interventions have the potential to have a positive impact on recruitment and retention of the rural medical workforce. **The WHO proposes a framework of six dimensions to guide the selection of appropriate bundles of interventions based on relevance, acceptability, feasibility, affordability, effectiveness and impact** (WHO 2021). These dimensions can then be used to measure development, attractiveness, recruitment and retention of the health workforce within rural and remote areas (WHO 2021).

Verma et al. (2016) reported that for GPs and GP trainees they were unable to draw any conclusions about retainer schemes, re-entry schemes, international recruitment, specialised recruiters, support for professional development or research, delayed partnerships, well-being or peer support or mixed approaches. This rapid review of reviews did not find any additional evidence for these interventions for GPs and GP trainees.

Whilst the majority of evidence found was related to the rural international context within USA, Canada or Australia, the principles behind the interventions can be transferred to the Welsh context, especially with regard to undergraduate and postgraduate education across the different healthcare disciplines.

### 3.2 Limitations of the available evidence

While the included reviews searched for literature related to the recruitment and retention of a wide range of healthcare professionals, such as doctors, dentists, nurses, and AHPs (including occupational therapists, physiotherapists, pharmacists, dietitians, clinical psychologists, and speech and language therapists), **limited evidence was found on interventions aimed at AHPs, nurses and dentists**. Moreover, the majority of the evidence on AHPs included studies looking at mixed professional groups and thus profession specific effects of the interventions cannot always be separated.

Many of the reviews mention that **finding a single, global definition for ’rural area’ was not possible, due to a lack of consistency or insufficient detail in how the primary studies described ’rural’** (Noya et al. 2021, Russell et al. 2021, WHO 2020, 2021). Moreover, national classifications and standards used to define ’rural area’ were often different within primary studies conducted in the same country, adding to the difficulty to find an all-encompassing definition (Noya et al. 2021).

There was very little high quality evidence presented across all the included reviews and the majority, consisted of a variety of moderate, low-quality and very low primary research studies as rated by the individual review authors. The majority used cohort or cross-sectional designs, which often lacked comparison groups, leading to uncertainty whether the change in recruitment and retention rates can be attributed to the intervention. Noya et al. (2021) comments that implementing randomised controlled trials (RCTs) in the real world to investigate recruitment and retention initiatives can be difficult as ethical issues around inequitable recruitment, forced training, and mandatory work for those not interested in rural training could arise. However, there are pragmatic RCT designs that can be used to overcome some of these challenges, and robust evaluations (or natural experiments) can also be designed and planned in advance of the rollout of any innovation. Another issue that we encountered and reported by the review authors was that many of the primary studies did not apply appropriate statistical analysis to confirm their findings.

### 3.3 Implications for policy and practice

The review **identifies a range of interventions that can be used for enhancing recruitment and retention of NHS clinical staff in Wales, particular in rural areas**. The evidence also supports the use of multiple-component interventions.

The findings **highlight the importance of providing and locating undergraduate and post graduate training in rural locations**. The findings also corroborate the use of bursary schemes for training, such as those already available for Nursing in Wales.

Further, more **robust evaluations, based on comparative studies, are required** to assess the effectiveness of interventions to support recruitment and retention of clinical staff. There was limited evidence on interventions aimed at allied health professionals.

### 3.4 Strengths and limitations of this Rapid Review

Limitations of this rapid review of reviews mainly originate from the issues with the available evidence identified in Section 2.2) which impact on the generalisability of the findings. In addition, the interventions mentioned in this rapid review are mainly based on cohort and descriptive studies and so we cannot be fully certain of the benefits. Further research is needed to determine the effectiveness of interventions for recruitment and retention of healthcare professionals, although certain ethical issues around forced training and labour need to be considered.

The strength of this review is that a thorough search was undertaken by an information specialist across five electronic databases, and the websites of 35 organisations were searched. Although this was a rapid review of reviews in which several of the systematic review processes could have been streamlined, it should be noted that data screening, data extraction and critical appraisal of each study were undertaken by different reviewers and then independently checked for accuracy and consistency by the same second reviewer.

The synthesis identified overall that there was reasonable agreement among all the included literature, which may be considered to imply some degree of reliability. However, findings can be profession and context dependent, which need to be considered when interpreting the results. Primary research studies focusing on interventions implemented in Europe and UK were separately summarised which is a strength of this review, as these strategies might be transferable to the Welsh context.

## Abbreviations

AHPs: Allied health professionals
GP: General Practitioner
GRADE: Grading of Recommendations Assessment, Development and Evaluation
HIC: High income country
IMGs: International medical graduates
LMIC: Low and middle income country
MIC: Middle income country
NHS: National Health Service
NHI: National Health Insurance
PCT: Primary care team
RCTs: Randomised controlled trials
RoS: Return of Service
WHO: World Health Organisation

## Data Availability

All data produced in the present work are contained in the manuscript

## 5. RAPID REVIEW METHODS

### 5.1 Eligibility criteria

The PICoS framework was used to inform the eligibility criteria: **P**opulation, Phenomena of **I**nterest, **Co**ntest and **S**tudy design. The population was informed by healthcare and education shortage occupations list (UK Visas and Immigration 2021).

**Table 3:**
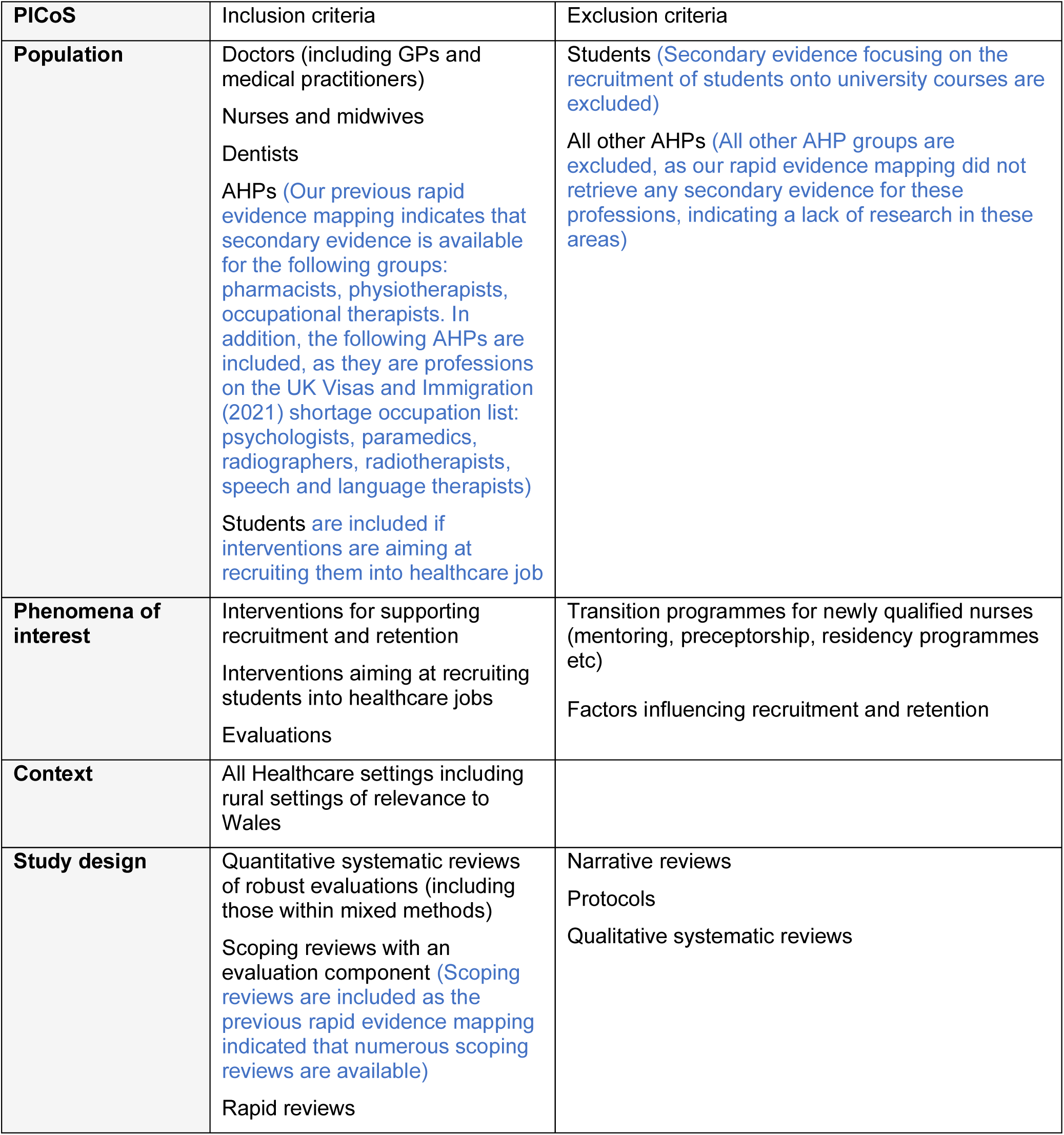
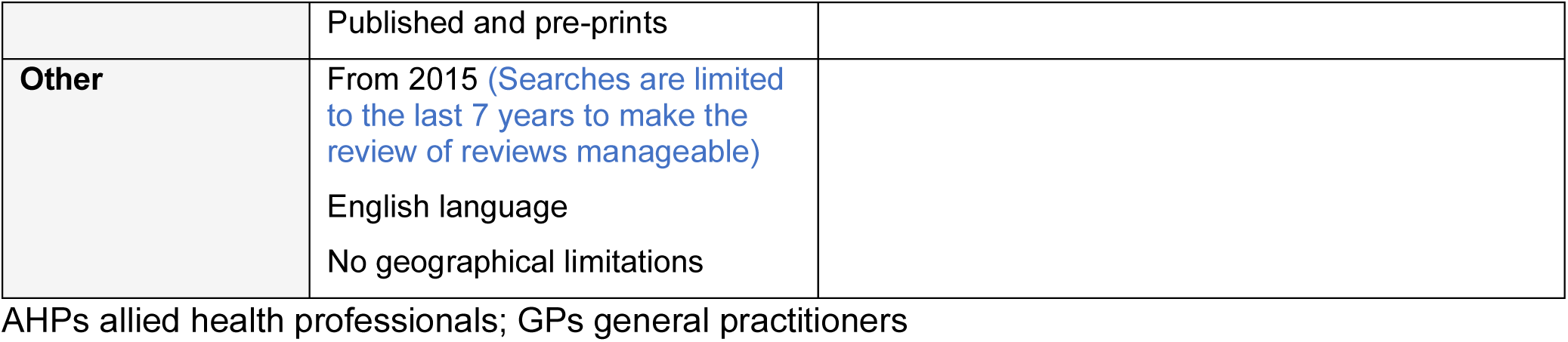
Eligibility Criteria.

### 5.2 Literature search

Comprehensive searches were conducted across five databases for English language publications from 2015 to February 2022:

- On the Ovid Platform: Medline, Embase, Emcare, HMIC
- On the Ebsco Platform: Cumulative Index of Nursing and Allied Health Literature
- Epistemonikos
- CENTRAL

The websites of key third sector and government organisations were also searched (information is in additional material, which is available on request).

An initial search of MEDLINE was undertaken (retention or retain* or recruit* AND doctor OR nurs OR midwi* OR dentist OR dental OR general practitioner* OR Pharmacist*, Physiotherapist*, occupational therapist*, paramedic*, radiographer*, radiotherapist* AND review* or meta*) followed by analysis of the text words contained in the title and abstract, and of the index terms used to describe article. This informed the development of a search strategy which was tailored for each information source. The full search strategies for each of the databases are provided (additional material is available on request). The existing umbrella reviews and rapid reviews that include reviews were used to identify relevant systematic reviews.

All citations retrieved from the database searches were imported into EndNote^TM^ (Thomson Reuters, CA, USA) and duplicates removed. Irrelevant citations were removed by searching for keywords within the title using the search feature within the Endnote software. The project team agreed which keywords to use to identify papers which did not meet the inclusion criteria. At the end of this process the citations that remained were exported as an XML file and then imported to Covidence^TM^.

### 5.3 Study selection process

Two reviewers dual screened all the citations using the information provided in the title and abstract using the software package Covidence^TM^, resolving all conflicts. For citations that appeared to meet the inclusion criteria, or in cases in which a definite decision could not be made based on the title and/or abstract alone, the full texts of all citations were retrieved. The full texts were screened for inclusion by two reviewers using the software package Covidence^TM^, a third reviewer was not required as there were no disagreements.

### 5.4 Data extraction

All demographic data was extracted directly into tables by one reviewer and checked by another. The data extracted included specific details about the populations, study methods and outcomes of significance to the review question and specific objectives. All outcome data were extracted by one reviewer and checked by a second.

### 5.5 Quality appraisal

Eligible systematic reviews were critically appraised using the JBI critical appraisal checklist for systematic reviews and research syntheses (Aromataris et al. 2015). All systematic regardless of the results of their methodological quality, underwent data extraction and synthesis (where possible). The results of the critical appraisal are reported in narrative form and in a table. Methodological quality assessment was conducted by one reviewer and checked by a second, there were no disagreements.

### 5.6 Data summary

The overlap of original research studies included in the systematic and rapid reviews was checked and reported in a table. To determine the degree of overlap, the corrected covered area (i.e. one primary study covered by multiple reviews) has been calculated (Pieper et al. 2014). Using this approach for the corrected covered area, less than 5% overlap is a slight overlap, 6-10% is a moderate overlap, 11-15% is a high overlap and >15% is a very high overlap. Seventy-four primary studies were duplicated across the systematic reviews (see additional material). The corrected covered score was found to be 3.1% (i.e., a slight overlap with systematic reviews mostly considering different primary studies). A total of 292 primary studies were cited by the included systematic reviews including 218 (74.6%) that were cited only once. All systematic reviews were included in this umbrella review regardless of the degree of overlap and percentage corrected covered area. The recency of the evidence base is as follows: 1970s (n=3); 1980s (n=1); 1990s (n=13), 2000s (n=275).

The data were reported narratively as a series of thematic summaries (Thomas et al. 2017) structured around the type of intervention, target population characteristics, type of outcome and intervention content. The type of intervention used the categories proposed by the WHO retention working group for their 2010 review on retention for healthcare workers which were educational interventions, regulatory interventions, financial incentives, personal and professional support and bundled interventions (activities that cover two or more different categories) (WHO 2010). Across all the included systematic reviews, some interventions were categorised differently, for example Verma et al. (2016) categorised bonded schemes as financial incentives, while Russell et al. (2021) considered these regulatory. Where there was a lack of consensus on the intervention categories, data was summarised in this rapid review of reviews according to the WHO framework (WHO 2010).

To gain further insight into interventions that could be transferable to the Welsh context, studies from the included systematic reviews and scoping reviews with evaluation components were filtered for UK and Europe-based primary research and a separate summary was reported. The earliest systematic review retrieved within our time frame was conducted by Grobler et al. (2015). This review sought to identify evidence for all qualified healthcare professionals which included doctors (GPs and specialists), nurses, occupational therapists, physiotherapists, speech and hearing therapists, pharmacists, dieticians, clinical psychologists and dentists. However, only one study was retrieved which was conducted in Thailand for Western medicine physicians, Chinese medicine physicians and dentists. The next earliest review was conducted by Verma et al. (2016) and this focused on GPs and covered a wide of range of educational interventions, regulatory interventions, financial incentives, personal and professional support and bundled interventions. The review was well conducted and on appraisal scored 9 out of 11 in the JBI checklist for systematic reviews and research syntheses and was written with the UK context in mind (32% of the included studies were conducted in European countries). It was therefore decided to use this review as a baseline and to use the primary research studies within the later systematic reviews to update the evidence base.

## 6. EVIDENCE

### 6.1 Study selection flow chart

The flow of citations through each stage of the review process is displayed in a PRISMA flowchart (Page et al. 2021), see Figure 1.

**Figure 1:**
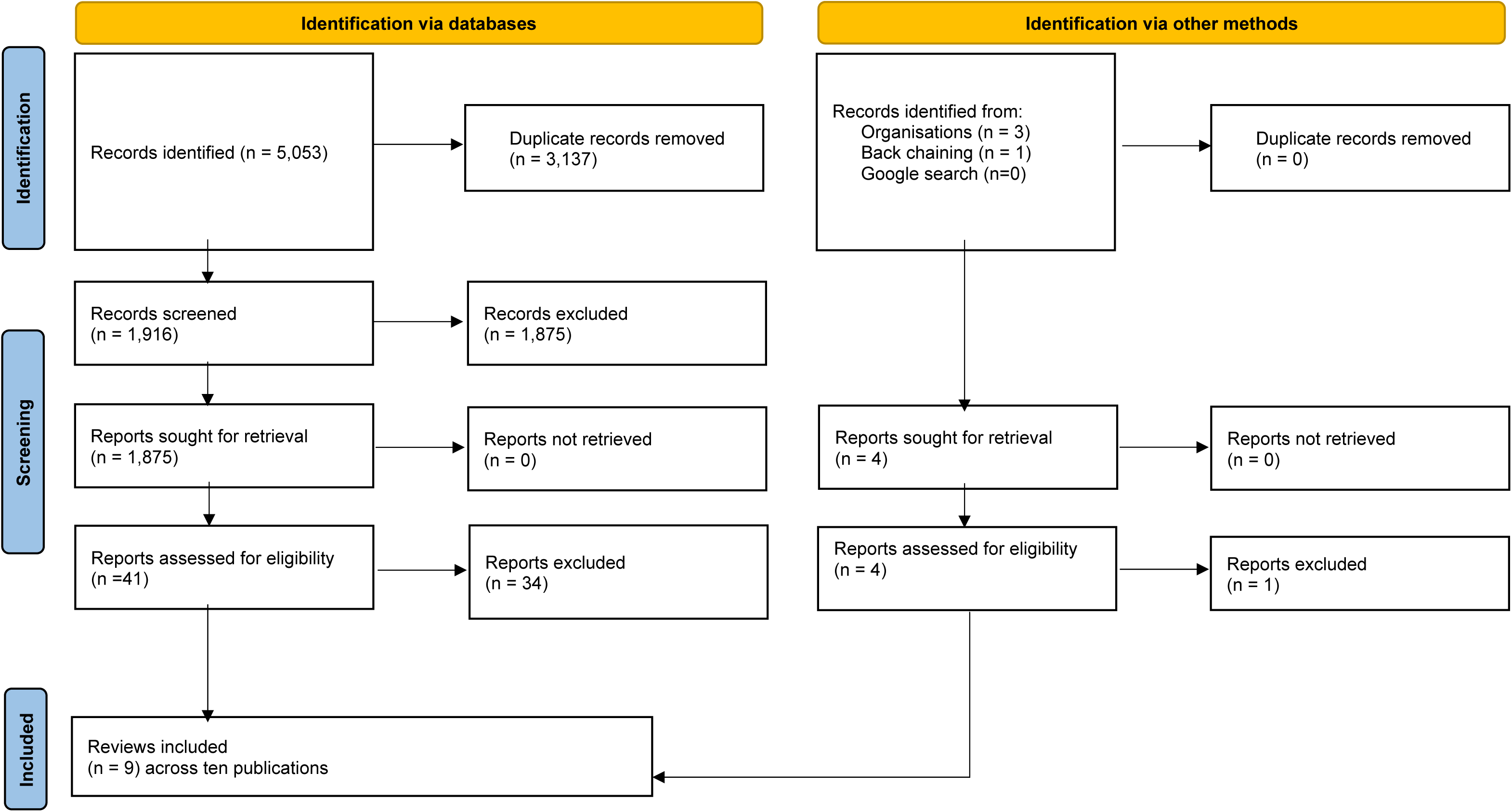
PRISMA 2020 flow diagram

### 6.2 Additional information available on request

1. Full search strategies
2. List of grey literature resources searched
3. Critical appraisal scores
4. Excluded studies
5. Protocol

## 7.1 Acknowledgements

The authors would like to thank Luke Davies, Charlette Middlemiss, Ian Owen and Sally Anstey for their contributions in guiding the focus of the review and in interpreting the findings.

## 8. ABOUT THE WALES COVID-19 EVIDENCE CENTRE (WCEC)

The WCEC integrates with worldwide efforts to synthesise and mobilise knowledge from research.

We operate with a core team as part of Health and Care Research Wales, are hosted in the [Wales Centre for Primary and Emergency Care Research (PRIME)](http://www.primecentre.wales/), and are led by [Professor Adrian Edwards of Cardiff University](https://www.cardiff.ac.uk/people/view/123022-edwards-adrian).

The core team of the centre works closely with collaborating partners in [Health Technology Wales](https://www.healthtechnology.wales/), Wales Centre for Evidence-Based Care, [Specialist Unit for Review Evidence centre](https://www.cardiff.ac.uk/specialist-unit-for-review-evidence), [SAIL Databank](https://healthandcareresearchwales.org/about-research-community/secure-anonymised-information-linkage-sail-databank), [Bangor Institute for Health & Medical Research/Health and Care Economics Cymru](https://www.bangor.ac.uk/health-sciences/research/index.php.en), and the [Public Health Wales Observatory](https://phw.nhs.wales/services-and-teams/observatory/).

Together we aim to provide around 50 reviews per year, answering the priority questions for policy and practice in Wales as we meet the demands of the pandemic and its impacts.

### Director

Professor Adrian Edwards

### Contact Email

**Website:** https://healthandcareresearchwales.org/about-research-community/wales-covid-19-evidence-centre

All reports can be downloaded from the library on the WCEC website.

## 9. APPENDICES

**Table 1:** Summary table of included systematic and scoping reviews.

| Citation | Review details | Included studies | Quality of the primary research<br>Quality of the review | Findings |
| --- | --- | --- | --- | --- |
| Grobler et al. 2015<br>https://doi.org/10.1002/14651858.cd005314.pub3<br>Systematic review | <u>Review period</u><br>Inception to 2015<br><u>Review purpose</u><br>To assess the effectiveness of interventions aimed at increasing the proportion of health professionals working in rural and other underserved areas<br><u>Included study designs</u><br>Cohort study (Interrupted trend analysis with time series observations) (n=1)<br><u>Outcomes of interest</u><br>Primary outcomes<br>The proportion of healthcare professionals who initially choose to work in rural or urban underserved areas as a consequence of being exposed to the intervention (recruitment)<br>The proportion of healthcare professionals who continue to work in rural or urban underserved areas as a consequence of the intervention (retention) | <u>Number of included studies</u> 1<br><u>Interventions</u><br>Educational interventions (n=0)<br>Regulatory interventions (n=1)<br>Financial incentives (n=0)<br>Personal and professional support (n=0)<br><u>Participants</u><br>Western medicine physicians, Chinese medicine physicians and dentists (n=1)<br><u>Countries of interventions</u><br>Taiwan (n=1)<br><u>Settings</u><br>Rural and underserved areas<sup>c</sup> | <u>Appraisal scale</u><br>Cochrane RoB for interrupted time series<br>GRADE<br><u>Appraisal rating</u><br>Serious concerns for risk of bias<br>Very low certainty of evidence<br><u>Review appraisal score</u><br>10 out of 11 in the JBI checklist for systematic reviews and research syntheses | Implementation of National Health Insurance scheme on equity of physician distribution<br>The equality of the geographic distribution of the health providers was measured using Gini coefficients (Gini coefficient of 0 = no inequality; Gini coefficient of 1 = complete inequality) for each of the 21 prefectures/cities in Taiwan. The investigators examined the distribution of health providers 24 years<br>*Equity of distribution of Western medicine physicians post implementation*<br>1 year post implementation (SE - 0.008, 0.004, p=0.071)<br>4 years post implementation (SE - 0.018, 0.003, p=0.00)<br>8 years post implementation (SE - 0.032, 0.005, p=0.00)<br>*Equity of distribution of Chinese medicine physicians post implementation*<br>1 year post implementation (SE - 0.007, 0.013, p=0.589) |
|  |  |  |  | 4 years post implementation (SE - 0.016, 0.011, p=0.172)<br>8 years post implementation (SE - 0.025, 0.016, p=0.143)<br>*Equity of distribution of dentists post implementation*<br>1 year post implementation (SE - 0.013, 0.017, p=0.444)<br>4 years post implementation (SE - 0.039, 0.03, p=0.196)<br>8 years post implementation (SE - 0.073, 0.056, p=0.201) |
| Johnson et al. 2018<br>https://doi.org/10.1186/s12909-018-1287-y<br>Systematic review | <u>Review period</u><br>January 2005 to January 2017<br><u>Review purpose</u><br>To examine the evidence base of rural educational programs within medical education and focuses on workforce intentions and employment outcomes<br><u>Included study designs</u><br>Cohort studies (n=39)<br>Descriptive studies (n=19)<br>Mixed methods study (n=1)<br>Qualitative studies (n=2)<br>Quasi-experimental studies (n=1)<br><u>Outcomes of interest</u><br>Future rural intentions<br>Working locations | <u>Number of included studies</u> 62<br><u>Interventions</u><br>Rural Clinical Placement Programs (n=24)<br>Rural Clinical Placement Programs combined with a rural health educational curriculum component (n=12)<br>Rural Clinical School Programs (n=23)<br>Studies comparing two of the above interventions (n=3)<br><u>Participants</u><br>Medical undergraduates<br><u>Countries of interventions</u><br>Australia (n=35), USA (n=18), Canada (n=5), New Zealand (n=2), Thailand (n=1) and Africa (n=1)<br><u>Settings</u> | <u>Appraisal scale</u><br>Modified Type of Evidence Schema<br>Modified Health Gains Notation Framework<br>Modified Cochrane Health Promotion and Public Health Field quality assessment screening questions for quantitative and qualitative studies<br><u>Appraisal rating</u><br>Strong (n=6)<br>Moderate (n=40)<br>Weak (n=16)<br><u>Review appraisal score</u><br>9 out of 11 in the JBI checklist for systematic reviews and research syntheses | There were mixed findings reported for rural placement or rural clinical school programs and rural intentions or actual graduate rural employment<br>Evidence from 34 studies (55%) provides strong evidence about the effectiveness of educational interventions<br>*Workforce outcomes*<br>43 of the 62 studies showed increased student interest (or increased association) of working in a rural location<br>28 out of 62 studies reported increased rural intentions or an increased likelihood to work rurally<br>22 out of 62 studies reported increased student interest in rural health medicine |
|  |  | Rural (no further details provided) |  | 7 out of 62 studies reported long term retention of graduates employed in a rural location<br>5 out of 62 studies did not report a positive association with graduates choosing rural employment<br>*Outcomes classified by interventions*<br>The Rural Clinical Placement Programs had the highest number of studies reporting a positive rural association (n=21/24, 88%) compared to (n=15/23, 65%) studies for the Rural Clinical School Programs |
| Kumar and Clancy 2020<br>https://doi.org/10.1093/pubmed/fdaa031<br>Systematic review | <u>Review period</u><br>Inception to 2019<br><u>Review purpose</u><br>To summarises the recent literature identifying factors that influence rural doctor retention and analyses strategies implemented to increase retention<br><u>Included study designs</u><br>Cohort studies (n=7)<br>Descriptive studies (n=7)<br>Systematic reviews (n=5)<br><u>Outcomes of interest</u><br>Proportion of doctors who chose to work in a rural area and the duration of this work | <u>Number of included studies</u> 19<br><u>Interventions</u><br>Educational interventions<br>Regulatory interventions<br>Financial incentives<br>Personal and professional support<br>Bundled strategies<br><u>Participants</u><br>Physicians<br><u>Countries of interventions</u><br>Africa (n=1), Australia (n=1), India (n=1), USA (n=10), Zambia (n=2), Multiple countries (n=4) | <u>Appraisal scale</u><br>Modified Newcastle-Ottawa scale<br>Tools by the National Institute of Health<br><u>Appraisal rating</u><br>Moderate (n=11)<br>High (n=8)<br><u>Review appraisal score</u><br>7 out of 11 in the JBI checklist for systematic reviews and research syntheses | HMICs<br>*Effective strategies*<br>Rural-based training programmes (elective or compulsory)<br>Sustainable workplace organisation and infrastructure<br>Social supports<br>*Non-effective strategies*<br>Visa waiver programs<br>*Inconclusive strategies*<br>Financial incentives<br>Although it was suggested that direct financial incentives without RoS are most effective, particularly loan repayment programs (in countries with high cost of medical education) |
|  |  | <u>Settings</u><br>Rural<sup>d</sup><br>Studies from countries of all income levels were included<sup>e</sup> |  | MICs<br>*Effective strategies*<br>Rural placements<br>Financial incentives<br>Workplace infrastructure<br>*Non-effective strategies*<br>Independently implemented strategies without consistency<br>*Inconclusive strategies*<br>Compulsory rural service programs<br>LMICs<br>A single study identified rural-based learning programs as a way to educate and incentivize health practitioners to work in rural locations<br>Implemented strategies must be multifactorial or bundled and relevant to the local context. |
| MacQueen et al. 2018<br>https://doi.org/10.1007/s11606-017-4210-z<br>Systematic review | <u>Review period</u><br>2005 to March 2017<br><u>Review purpose</u><br>To assess reasons for current providers' geographic choices and the success of training programs aimed at increasing rural provider recruitment<br><u>Included study designs</u><br>Cohort studies (n=24) | <u>Number of included studies</u> 24<br><u>Interventions</u><br>Educational interventions<br>- Rural training programmes (n=24)<br><u>Participants</u><br>Medical students (n=11)<br>Medical resident training (n=11)<br>Nurse practitioners (n=1) | <u>Appraisal scale</u><br>Critical appraisal concentrated on the representativeness of the sample (selection bias), the response or follow-up rate (attrition bias), the role of confounding variables (e.g., lack of multivariate analyses), and the data source reporting and | Based on the variation in estimates and the frequent presence of selection bias, we conclude that there is moderate quality evidence that the success rate of rural training programs ranges from 30 to 65%, and, on average, only one in two trainees is likely to enter rural care |
|  | <u>Outcomes of interest</u><br>Practicing in rural care | Physician assistants (n=1)<br><u>Countries of interventions</u><br>USA (n=24)<br><u>Settings</u><br>Rural (as defined by the author)<br>US healthcare settings | reliability (detection bias). GRADE was also used.<br><u>Appraisal rating</u><br>Selection bias: high risk (n=12); low risk (n=7); unclear (n=4); N/A (n=1)<br>Attrition bias: high risk (n=1); low risk (n=2); unclear (n=4); N/A (n=17)<br>Confounding variables: low risk (n=5); unclear (n=1); N/A (n=18)<br><u>Review appraisal score</u><br>8 out of 11 in the JBI checklist for systematic reviews and research syntheses |  |
| Noya et al. 2021<br>https://dx.doi.org/10.34172/ijhpm.2021.160<br>Scoping review with evaluation component | <u>Review period</u><br>Jan 2010 to Nov 2020<br><u>Review purpose</u><br>1. What factors have been shown associated with improved recruitment, development, and retention?<br>2. What strategies/approaches have been implemented to improve recruitment, development, and retention?<br>3. What is the evidence of the success of these approaches?<br>4. What are the similarities and differences between approaches implemented in HICs and LMICs?<br><u>Included study designs</u> | <u>Number of included studies</u> 62<br><u>Interventions</u><br>Educational interventions<br>Regulatory interventions<br>Bundled strategies<br><u>Participants</u><br>Medical workforce<br><u>Countries of interventions</u><br>Australia (n=23), Brazil (n=1), Canada (n=9), Chile (n=1), France (n=1), India (n=1), Japan (n=1), New Zealand (n=1), | <u>Appraisal scale</u><br>N/A<br><u>Appraisal rating</u><br>N/A<br><u>Review appraisal score</u><br>6 out of 11 in the JBI checklist for systematic reviews and research syntheses | We found that the strategies positively impacting recruitment, retention, and development of the rural medical workforce were educational, regulatory, financial incentives, and bundled strategies<br>This evidence is strong across international contexts, with significant probabilities and a higher likelihood of rural practice<br>There are similarities and differences between approaches implemented in HICs and LMICs<br>Educational interventions |
|  | Mixed methods studies (n=8)<br>Qualitative studies (n=3)<br>Cohort studies (n=20)<br>Descriptive surveys (n=22)<br>Quantitative studies design not specified (n=8)<br>Not specified (n=1)<br><u>Outcomes of interest</u><br>Included but was not limited to outcomes for recruitment (rural and remote practice location; for development – personal and professional development); and for retention (continuity in rural and remote practice and low turnover rates). | Norway (n=3), Philippines (n=3), Thailand (n=3), UK (n=1), USA (n=14)<br><u>Settings</u><br>Rural and remote areas in HICs and LMICs (regardless the definition of rurality and rural background used in one country). |  | Most educational strategies internationally were implemented at the medical school-university level. These include rural background student selection, rural exposure during medical school, and rural oriented medical school<br>The undergraduate educational strategy was the most commonly reported strategy with positive results, though postgraduate training was also found to have significant results<br>Regulatory interventions<br>Recruiting IMGs has been one government level strategy that proved to be effective in rural medical workforce recruitment<br>More coercive strategies reported such as bonded scholarships and obligatory time commitment in rural areas have been met with some success<br>Bundled strategies<br>Chile and Thailand's bundled strategies provide good examples of what can be achieved with a holistic approach. Although there are some differences in terms of the target of the programs |
| Russell et al. 2021<br>https://doi.org/10.1186/s12960-021-00643-7 | <u>Review period</u><br>2010 to July 2020 | <u>Number of included studies</u> 34<br><u>Interventions</u> | <u>Appraisal scale</u> | Educational Interventions<br>There is growing evidence that preferential selection of students |
| Systematic review | <u>Review purpose</u><br>To examine the quantifiable associations between interventions to **retain** health workers in rural and remote areas of high-income countries and workforce retention<br><u>Included study designs</u><br>Cohort studies (n=34)<br><u>Outcomes of interest</u><br>Primary outcomes<br>Mean or median length of employment; survival probabilities; hazard, odds or relative risk ratios for staying/leaving rural; stability rates; settlement rates. (profile = retention in rural/remote area or community)<br>Secondary outcomes<br>Vacancy rates; unfilled positions; turnover numbers or rates; attrition or wastage rates; rate of leaving before | Educational interventions (n=21)<br>Regulatory interventions (n=9)<br>Financial incentives (n=5)<br>Personal and professional support (n=4)<br>Health systems (n=1)<br><u>Participants</u><br>Doctors (n=28), nurses (n=3), dentists (n=1), mixed groups of health professionals (n=2)<br><u>Countries of interventions</u><br>Australia (n=13), USA (n=11), Canada (n=5), Norway (n=4) and northern European countries of Iceland, Ireland, Norway, Scotland, Sweden, Greenland (n=1)<br><u>Settings</u><br>Rural or remote<sup>b</sup><br>High-income countries as per World Bank criteria or if mixed income countries, then data for high-income countries reported separately | JBI critical appraisal checklist for analytical cross-sectional studies<br>JBI critical appraisal checklist for cohort studies<br>JBI critical appraisal checklist for case reports<br><u>Appraisal rating</u><br>Methodological quality of included studies was generally low<br>The median score for included cohort studies was 16 out of a maximum of 22 (interquartile range 13–20)<br>Only one-third of included studies applied appropriate statistical analysis, with less than half adjusting for key potential confounders<br>Many studies had no comparator group<br><u>Review appraisal score</u><br>9 out of 11 in the JBI checklist for systematic | who grew up in a rural area is associated with increased rural retention. Undertaking substantial lengths of rural training during basic university training or during post-graduate training were each associated with higher rural retention, as was supporting existing rural health professionals to extend their skills or upgrade their qualifications<br>Regulatory interventions<br>Regulatory interventions requiring RoS in a rural area in exchange for visa waivers, access to professional licenses or provider numbers were associated with comparatively low rural retention, especially once the RoS period was complete. Rural retention was higher if RoS was in exchange for loan repayments<br>Financial incentives<br>Evidence about the impact of financial incentives (with no RoS requirement) was limited because of the small number of studies and failure to quantify actual retention behaviour of health professionals<br>Personal and professional support<br>Interventions such as, early sign up to internships, cognitive behavioural coaching programme, and enhanced professional network were identified in the literature, although the studies |
|  |  |  | reviews and research syntheses | were low quality, as due to the lack of adjustment for confounding factors, and comparator groups. One included study mentioned factors, such as reasonable hours, availability of locums and professional backup, and educational opportunities for children that could help retain professionals<br>Health systems<br>In the USA, expansion of Medicaid and Medicare in rural areas was associated with physicians moving away from the rural country and becoming clinically inactive |
| Suphanchaimat et al. 2016<br>https://dx.doi.org/10.2147%2FAMEP.S116699<br>Systematic review | <u>Review period</u><br>2000 to 2015<br><u>Review purpose</u><br>To assess the impact of strategies on the intention of dental students/graduates to practice in rural area<br><u>Included study designs</u><br>Descriptive surveys (n=6)<br>Retrospective cohort study (n=1)<br><u>Outcomes of interest</u><br>Event (providing dental services in rural areas, or where they showed a willingness/intention to practice in rural areas after graduation)<br>Non-events (that they were neither practicing in rural areas, nor had they | <u>Number of included studies</u> 7<br><u>Interventions</u><br>Educational interventions<br>- Rural exposure strategies<sup>a</sup> (n=7)<br><u>Participants</u><br>Dental students or dental graduates<br><u>Countries of interventions</u><br>USA (n=3), Australia (n=2), South Africa (n=1), Thailand (n=1)<br><u>Settings</u><br>Rural (including but not limited to remote areas, underserved areas, charitable works servicing underserved | <u>Appraisal scale</u><br>No tools used. The risk of bias and methodological limitations of the selected articles were described qualitatively in the data-extraction table<br><u>Appraisal rating</u><br>No quality rating was given to the primary studies. Quality issues included misuse of terminology to describe exposure and event; recall bias; selection bias; and low response rate among others<br><u>Review appraisal score</u> | Meta-analysis of all seven studies with random-effect model, showing results in terms of OR OR 4.06 (95% CI, 2.55–6.45) I<sup>2</sup>=66%<br>Meta-analysis of all seven studies with random-effect model, showing results in terms of RR RR 1.95 (95% CI, 1.37–2.77) I<sup>2</sup>=92.1%<br>Subgroup meta-analysis between rural background and rural clinical rotation<br>*Teaching programs concerning rural experience*<br>OR 4.32 (95% CI, 1.93-9.66) I<sup>2</sup>=64.7%<br>*Rural background* |
|  | demonstrated a willingness/ intention to do so) | populations, and community-based settings) | 6 out of 11 in the JBI checklist for systematic reviews and research syntheses | OR 4.20 (95% CI, 2.22–7.96)<br>I<sup>2</sup>=59.4% |
| Verma et al. 2016<br>https://doi.org/10.1186/s12913-016-1370-1<br>Systematic review | <u>Review period</u><br>Inception to Jan 2015<br><u>Review purpose</u><br>To evaluate interventions and strategies used to **recruit and retain** primary care doctors internationally<br><u>Included study designs</u><br>Descriptive surveys (n=38)<br>Cohort studies (n=12)<br>Pre Post test (n=1)<br><u>Outcomes of interest</u><br>Primary outcome<br>Number of primary care doctors recruited or retained<br>Secondary outcomes<br>Average duration of employment after recruitment<br>Future intentions | <u>Number of included studies</u> 51<br><u>Interventions</u><br>Financial incentives with RoS (n=11)<br>Recruiting rural students (n=6)<br>International recruitment (n=4)<br>Rural or primary care focused undergraduate placements (n=3)<br>Rural or underserved postgraduate training (n=3)<br>Wellbeing or peer support initiatives (n=3)<br>Marketing (n=2)<br>Mixed interventions (n=5)<br>Support for professional development or research (n=5)<br>Retainer schemes (n=4)<br>Re-entry schemes (n=1)<br>Specialised recruiters or case managers (n=2)<br>Delayed partnerships (n=2)<br><u>Participants</u><br>Primary care doctors<br>Studies that included medical specialties other than primary care were included if judged to be transferable to primary care<br><u>Countries of interventions</u><br>USA (n=18), UK (n=12), Australia (n=8), Canada (n=5), | <u>Appraisal scale</u><br>Modified Newcastle-Ottawa Scale<br><u>Appraisal rating</u><br>The studies were all of low methodological quality, and only 15 of the 51 included studies involved a comparison group<br>The representativeness of the included participants was generally good, however the absence of a comparison group resulted in a high risk of bias in many studies<br>Assessment of the outcome and follow-up was generally low risk of bias<br>Most studies were described in an adequate or detailed manner and had potential or good generalizability<br><u>Review appraisal score</u> | There is weak evidence from the 15 studies that included a comparison group that improved recruitment of primary care doctors was associated with **postgraduate placements in underserved areas, undergraduate rural placements and recruiting students to medical school from rural areas**<br>There was weak mixed evidence about **financial incentives with RoS**<br>The quality of the studies was not sufficient to draw conclusions about retainer schemes, re-entry schemes, international recruitment, specialised recruiters, support for professional development or research, delayed partnerships, well-being or peer support or mixed approaches |
|  |  | Norway (n=4), Japan (n=2), New Zealand (n=1), Chile (n=1)<br><u>Settings</u><br>Primary care and HICs (as defined by the OECD) | 9 out of 11 in the JBI checklist for systematic reviews and research syntheses |  |
| WHO 2020<br>https://apps.who.int/iris/handle/10665/337300<br>Systematic review<br>WHO 2021<br>https://apps.who.int/iris/bitstream/handle/10665/341139/9789240024229-eng.pdf<br>Systematic review and guideline | <u>Review period</u><br>Jan 2010 to end of Nov 2019<br><u>Review purpose</u><br>Evaluations of interventions which increase the availability of health practitioners in rural and remote areas across the world<br><u>Included study designs</u><br>Audits (n=2)<br>Case studies (n=2)<br>Cohort studies (n=20)<br>Descriptive surveys (n=20)<br>Mixed methods studies (n=25)<br>Pre post tests (n=5)<br>Program evaluations (n=2)<br>Qualitative studies (n=26)<br>Quasi-experimental studies (n=3)<br>Systematic review (n=1)<br><u>Outcomes of interest</u><br>• Increase in total number of health workers recruited; Decrease in vacancy rates; Increase in the number of disciplines and types of health workers (Recruitment: improved rural health worker recruitment)<br>• Reduction in turnover and unstable staffing (Retention: improved rural health worker retention) | <u>Number of included studies</u><br>106<br><u>Interventions</u><br>Educational interventions (n=40)<br>Regulatory interventions (n=23)<br>Financial incentives (n=12)<br>Personal and professional support (n=11)<br>Bundled strategies (n=20)<br><u>Participants</u><br>A comprehensive range of health professional and community health worker occupations (volunteer and paid) were identified. These included students (at different stages), recent graduates, senior practitioners and sub-specialist varieties or different qualification groups of the same profession (e.g. enrolled and registered nurses)<br><u>Countries of interventions</u><br>Afghanistan (n=1), Australia (n=24), Bangladesh (n=3), Brazil (n=5), Burkina Faso | <u>Appraisal scale</u><br>GRADE<br><u>Appraisal rating</u><br>Low (n=42)<br>Very low (n=33)<br>Moderate (n=31)<br><u>Review appraisal score</u><br>9 out of 11 in the JBI checklist for systematic reviews and research syntheses | Extracted from WHO 2021<br>**Interventions should be interconnected, bundled and tailored to the local context**<br>**Educational interventions**<br>*Enrolling rural background students in health worker education programmes*<br>Admitting students with a rural background can have a large positive effect on the availability of rural health workers. (GRADE: moderate)<br>*Locating health worker education facilities outside capitals and major cities*<br>Locating health training institutions in rural areas significantly improves the recruitment and retention of their graduates – evidence mainly from HICs (GRADE: low)<br>*Exposing students to rural and remote communities and rural clinical experiences*<br>Evidence from six studies, mainly from high-income countries, suggests that exposure of |
|  |  | (n=1), Cambodia (n=1), Canada (n=10), Chile (n=1), China (n=3), France (n=1), Germany (n=1), Ghana (n=4), India (n=2), Indonesia (n=2), Israel (n=1), Kenya (n=1), Liberia (n=1), Mali (n=1), Nepal (n=1), Norway (n=3), Pakistan (n=2), Philippines (n=2), South Africa (n=3), Thailand (n=4), Uganda (n=2), UK (n=2), United Republic of Tanzania (n=2), USA (n=15), Zambia (n=2), multiple countries (n=5)<br><u>Settings</u><br>Rural and remote areas<sup>f</sup> |  | students to rural community experiences and clinical rotations is significantly positively associated with recruitment and retention as rural health workers (GRADE: low)<br>*Exposure to rural health topics in health worker education.*<br>The two observational studies on medical students and doctors found that a rural-focused curriculum led to regional increases in rural health workforce densities of graduates from the institution (GRADE – low)<br>*Continuing professional development for health workers practising in rural communities*<br>Evidence from five observational studies on diverse health workers demonstrates a significant positive effect of continuing professional development on recruitment and retention in four out of the five. Although the fifth study did not find a statistically significant relationship between perceptions of continuing education and anticipated retention, it was found to statistically significantly improve job satisfaction (GRADE – low)<br>**Regulatory interventions**<br>*Evidence on enhanced scope<sup>g</sup> of practice in rural areas* |
|  |  |  |  | Four observational studies on different health worker occupation groups in high-income countries suggest that an enhanced scope of practice increases job satisfaction which positively influences retention and improves access to health care for rural communities with shortages of high-level health workers or specialists (GRADE – low)<br>*Introduction of different types of health workers with the appropriate training in rural and remote areas*<br>Evidence from five observational studies involving different types of health workers across different countries shows that the introduction of different types of health workers with the appropriate training in rural and remote areas boosts the density of health workers. All five studies reported this positive effect (GRADE – low)<br>Compulsory service<br>*Evidence from three* Observational studies focusing mainly on doctors and dentists suggests that the impact of compulsory service on retention is influenced by personal and professional factors (GRADE – low) The certainty of evidence for this intervention is low and the evidence on the effects of<br>compulsory service on the retention of participants remains limited.<br>*Agreements for return of service*<br>Evidence from four observational studies in mostly high-income countries demonstrated positive influences on the increase in the availability of service-obligated health workers through high rates of completion of the service agreements and varied retention rates (GRADE – low)<br>**Financial incentives**<br>*Incentives*<br>Eight observational studies suggest that incentives are positively linked to health workers' decisions to locate in (or remain in) a rural area. Four observational studies demonstrated that financial incentives had a moderate positive effect on the recruitment and retention of health workers in rural areas in both the short and medium term. However, other studies suggest that the observed effect may not have been large enough to achieve the desired effect on availability (GRADE – low)<br>**Personal and professional support**<br>*Investing in rural and remote infrastructure and services to ensure decent living conditions* |
|  |  |  |  | One mixed-methods study on factors that enhance long-term retention of community health workers suggests that the favourability of living conditions, most notably ease of transportation and stronger partnerships with the local health unit, has a positive effect<br>*Safe and secure working environment for health workers in rural and remote areas*<br>Evidence from three observational studies from high-income countries and from low- and middle-income countries suggests that policies that improve safety and security have the positive effect of lowering turnover and attrition rates (GRADE – low)<br>*Decent work*<br>Evidence from two observational studies from high-income countries and from low- and middle-income countries suggests that policies that improve working conditions and environment have the positive effect of lowering turnover and attrition rates (GRADE – low)<br>*Health workforce support networks for health workers in remote and rural areas*<br>Evidence from seven observational studies suggest that the availability of telehealth,<br>mobile health and electronic health to rural health workers would have a positive impact on their job performance and aid retention (GRADE – low)<br>*Career development and advancement programmes, and career pathways for health workers in rural and remote areas*<br>Evidence from four observational studies across different socioeconomic classes of countries suggests that having career development posts in rural areas will increase attractiveness and job satisfaction, which is associated with decreased turnover intention (GRADE – low)<br>*Development of networks, associations, and journals for health workers in remote and rural areas*<br>Evidence from the before-and after and cross-sectional study on physicians suggests that the development of rural professional networks can improve retention (GRADE – low)<br>Social recognition measures for health workers in remote and rural areas 17 observational studies on the impact of factors that lead to retention of rural health workers in developing countries ranked recognition by the employer and community as being one of the<br>vital motivating factors (GRADE – very low) |

**Table 2:**
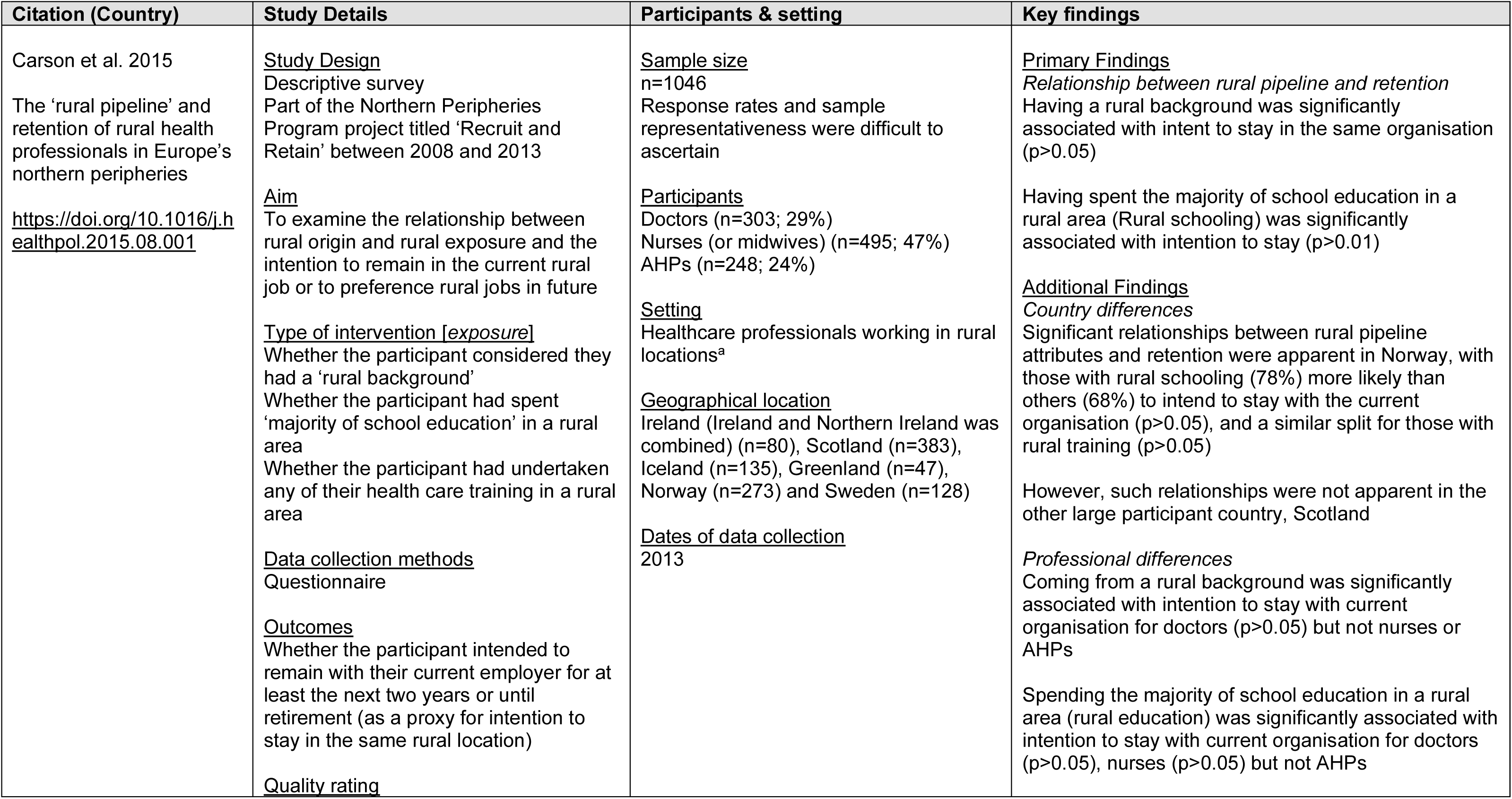
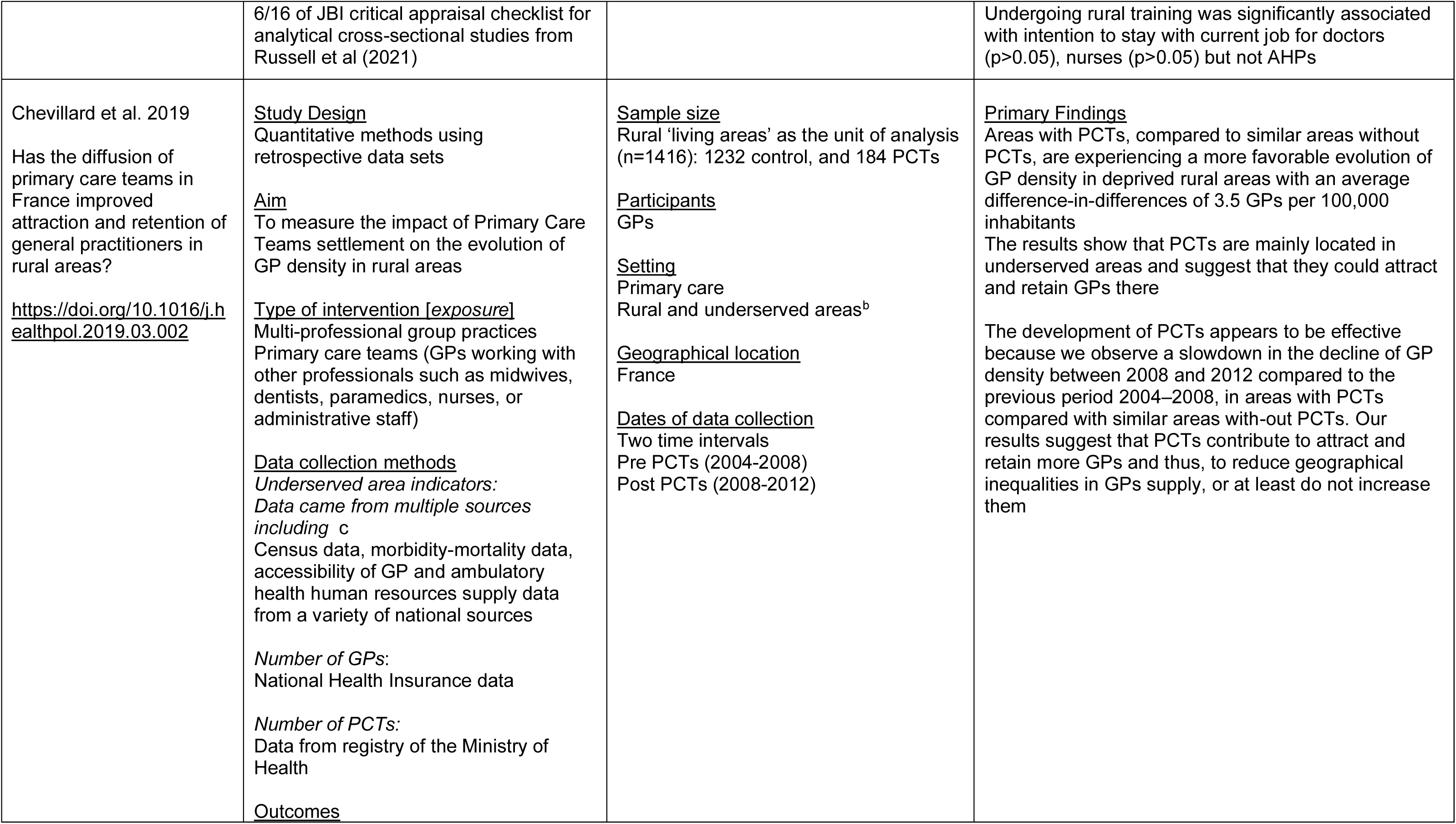
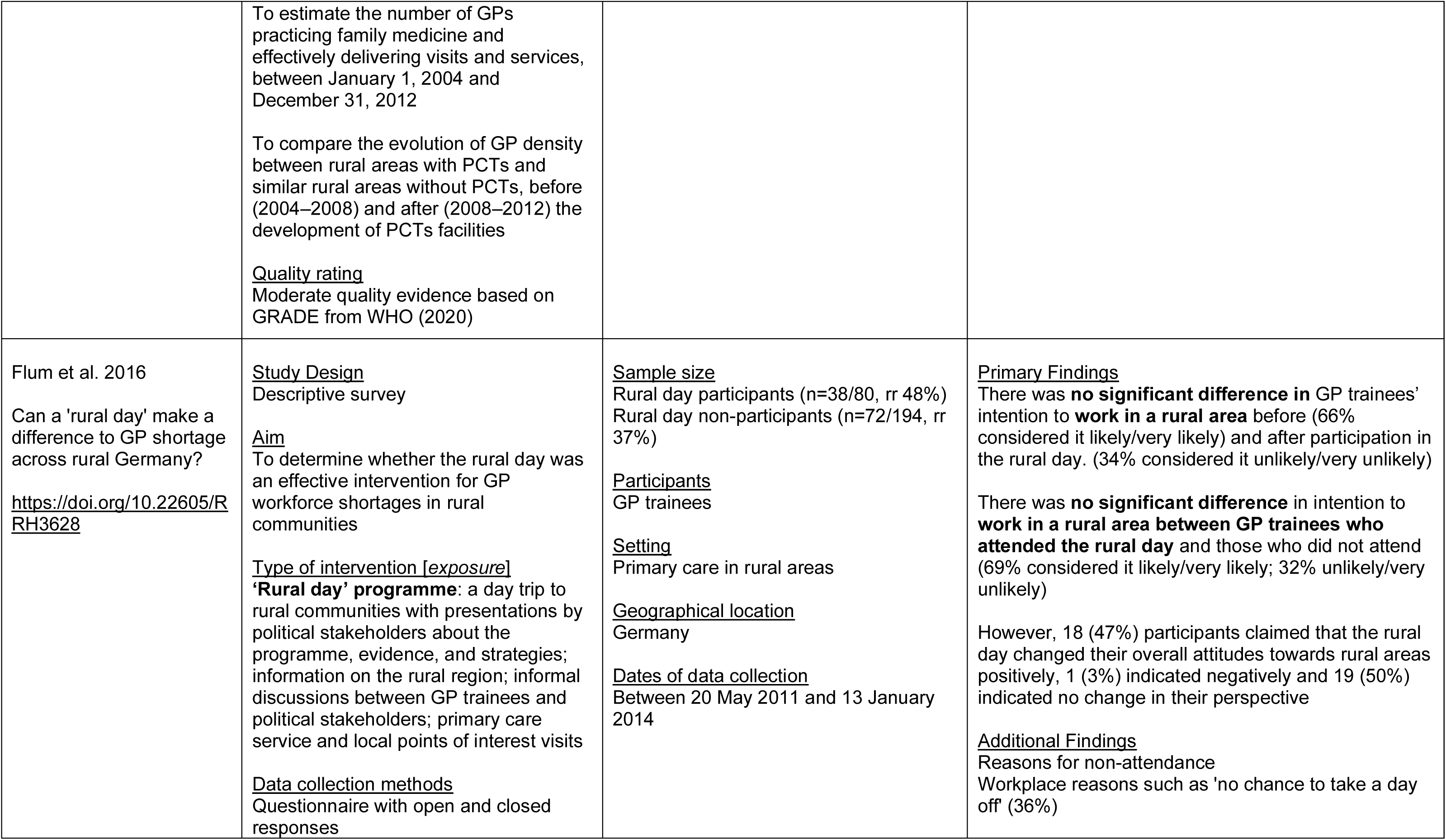
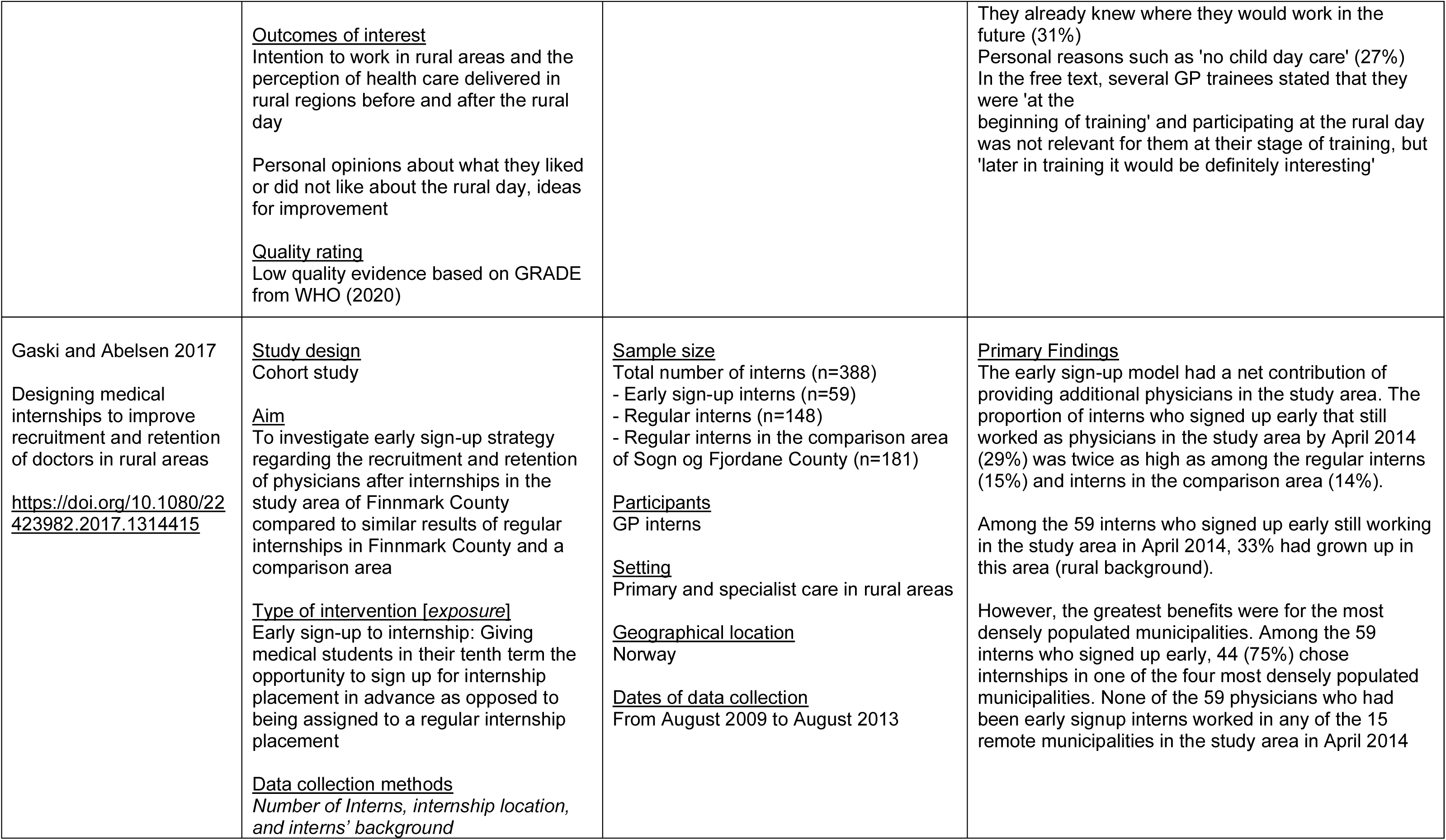
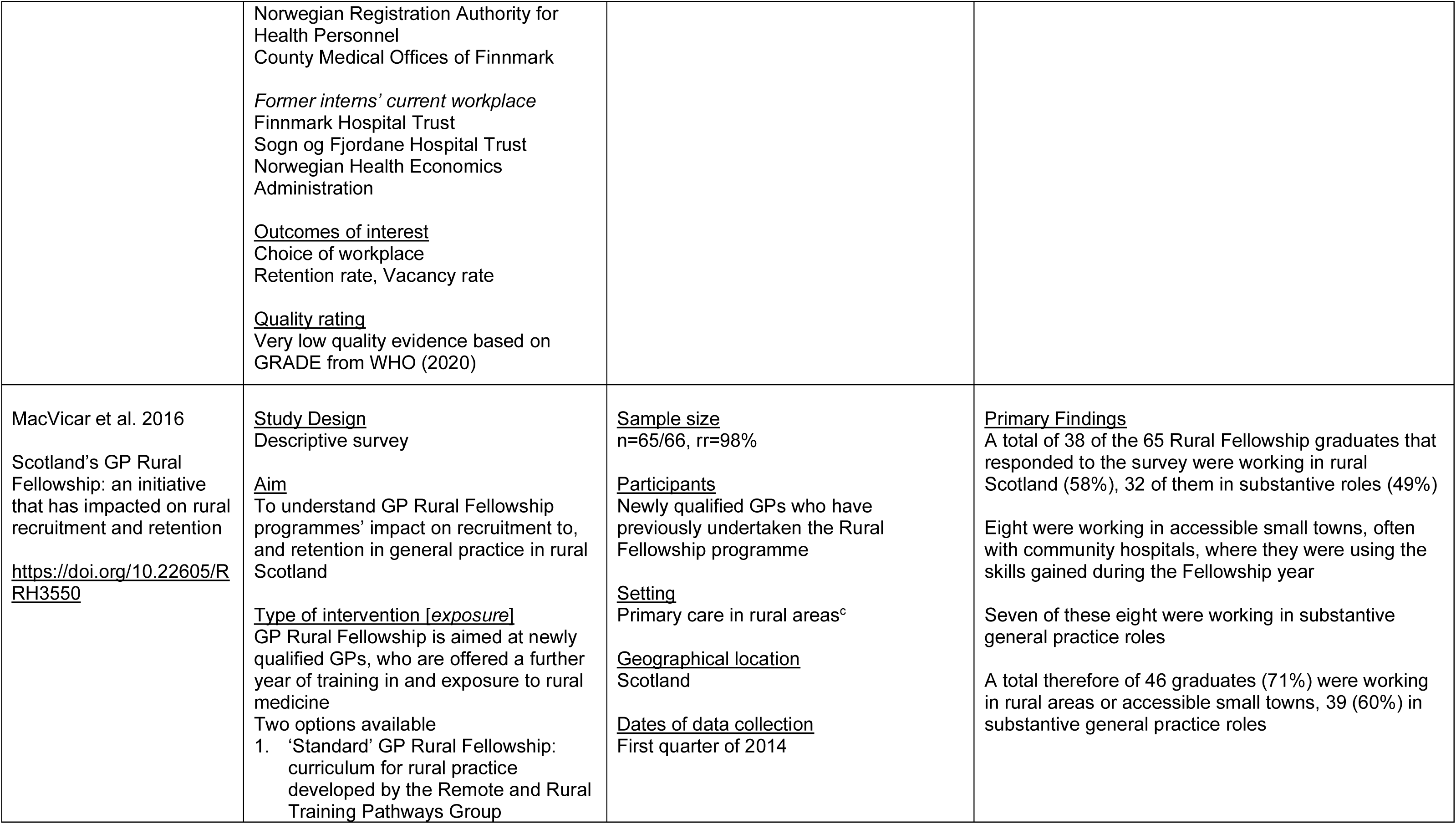
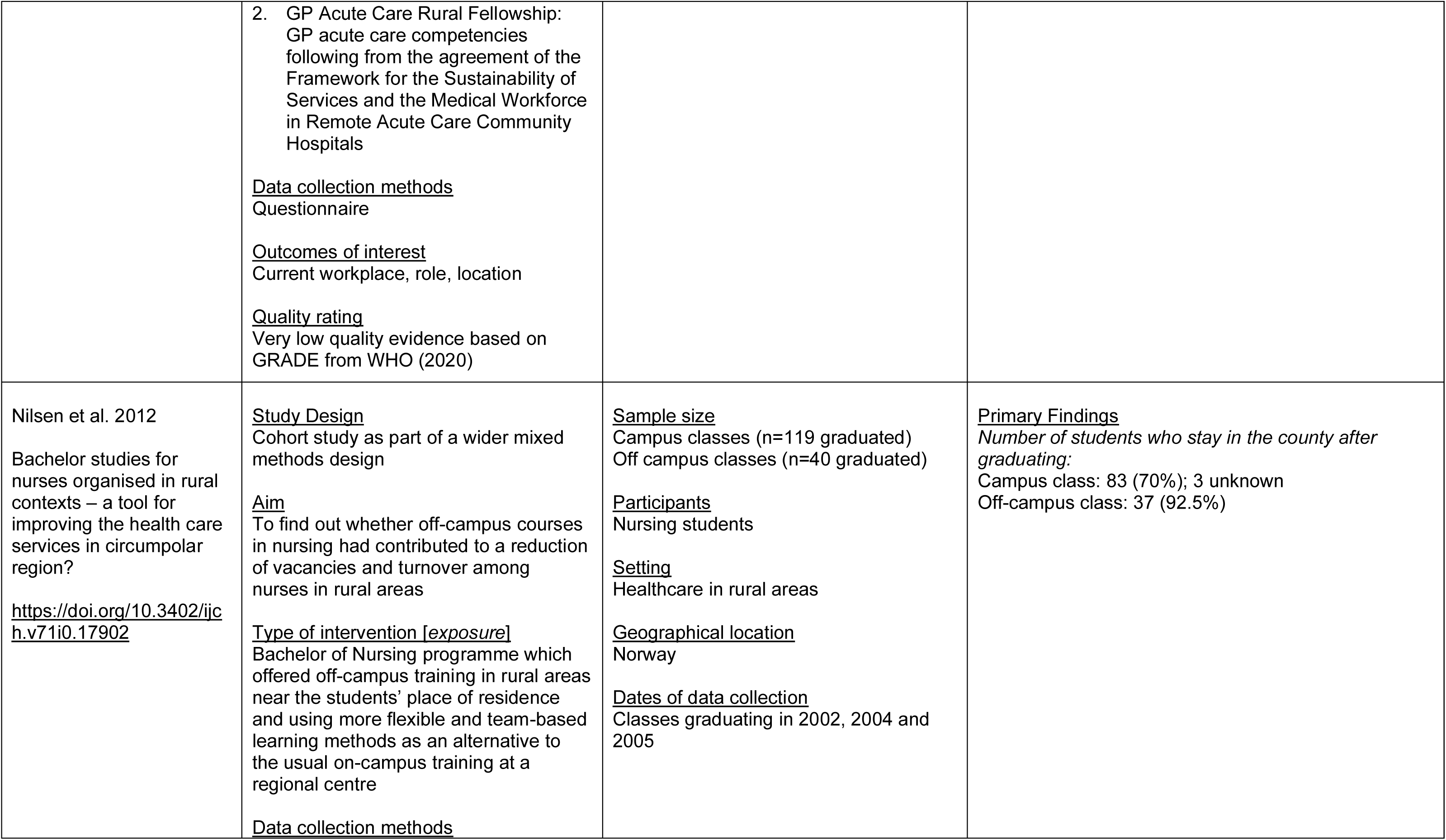
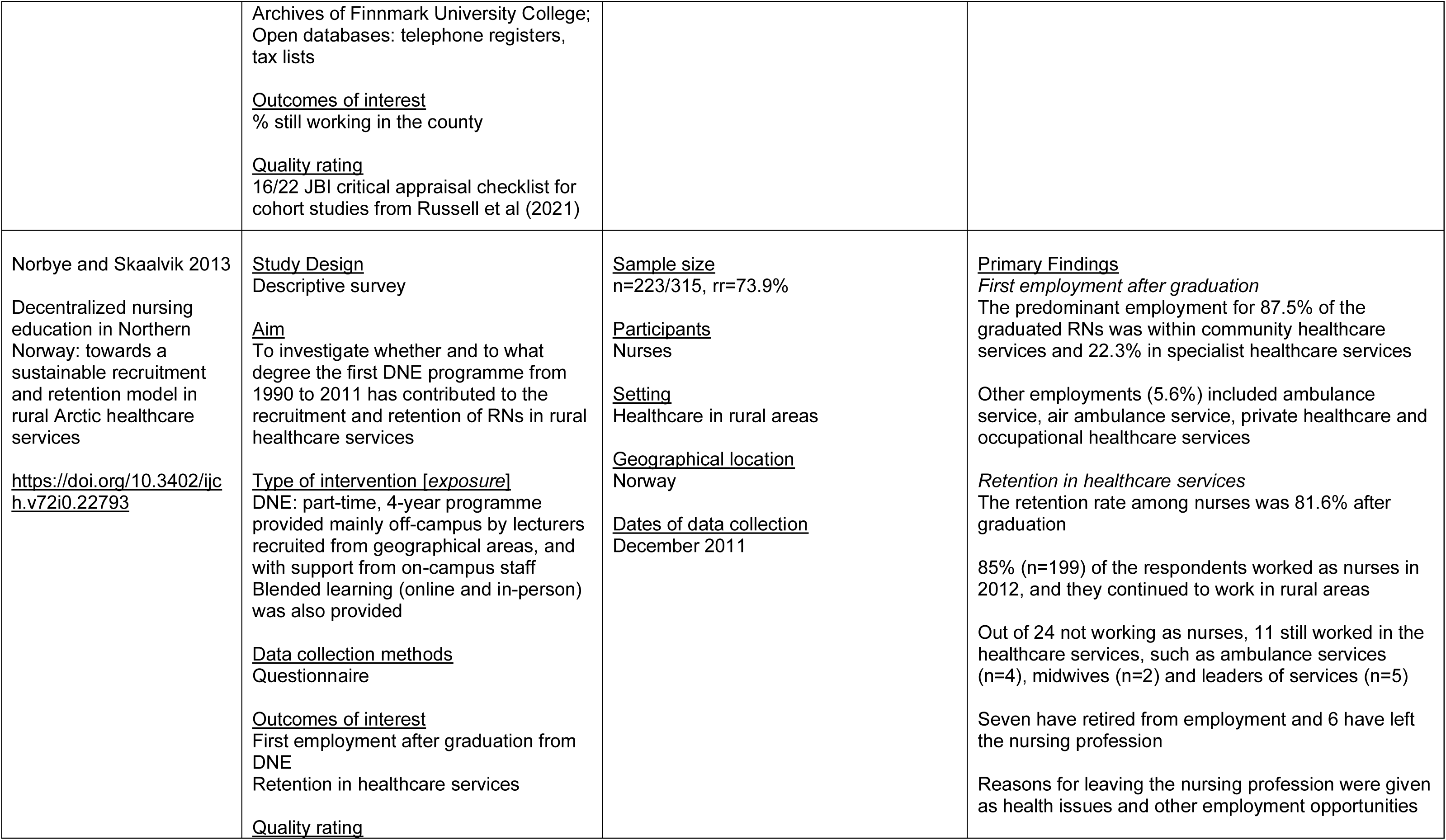
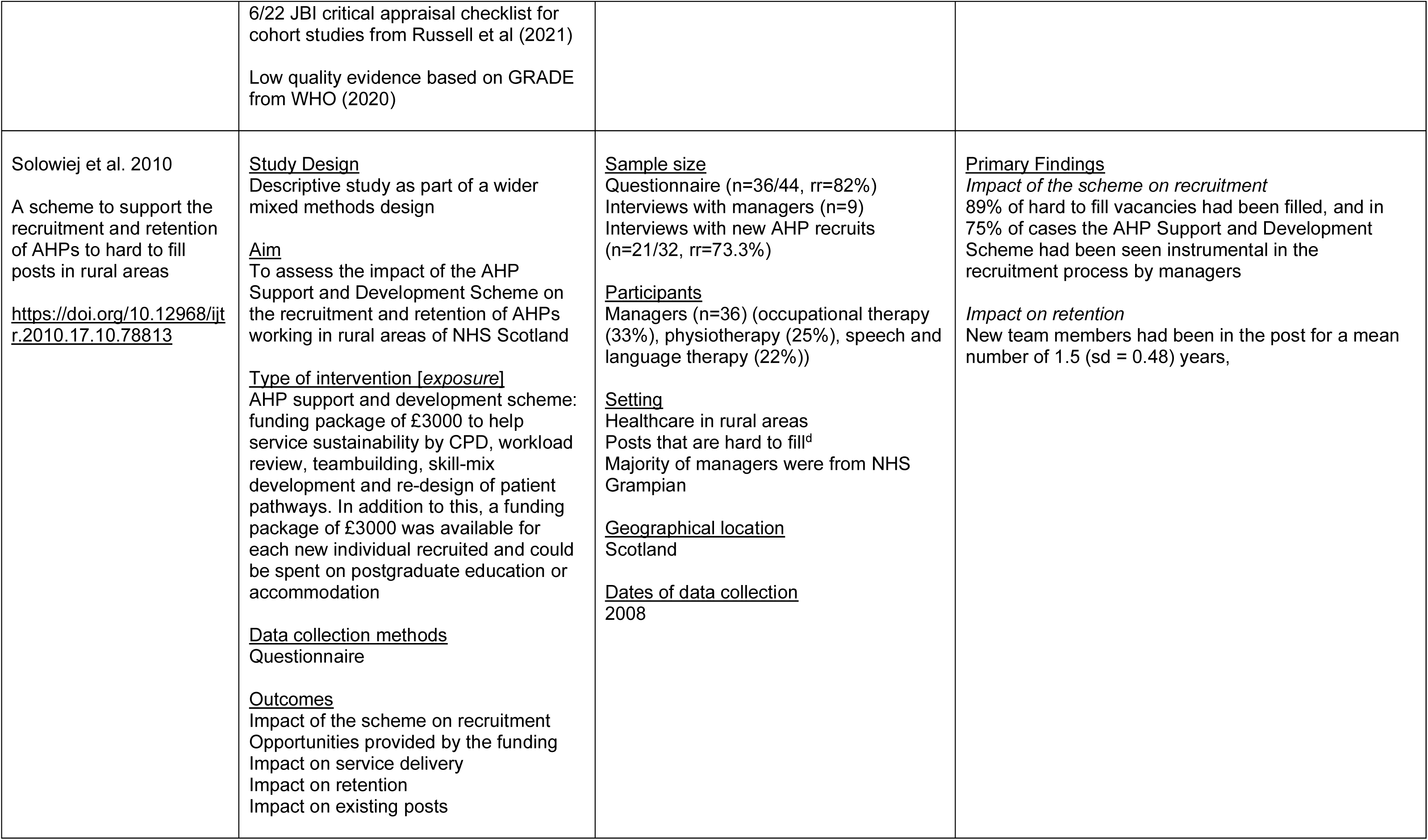
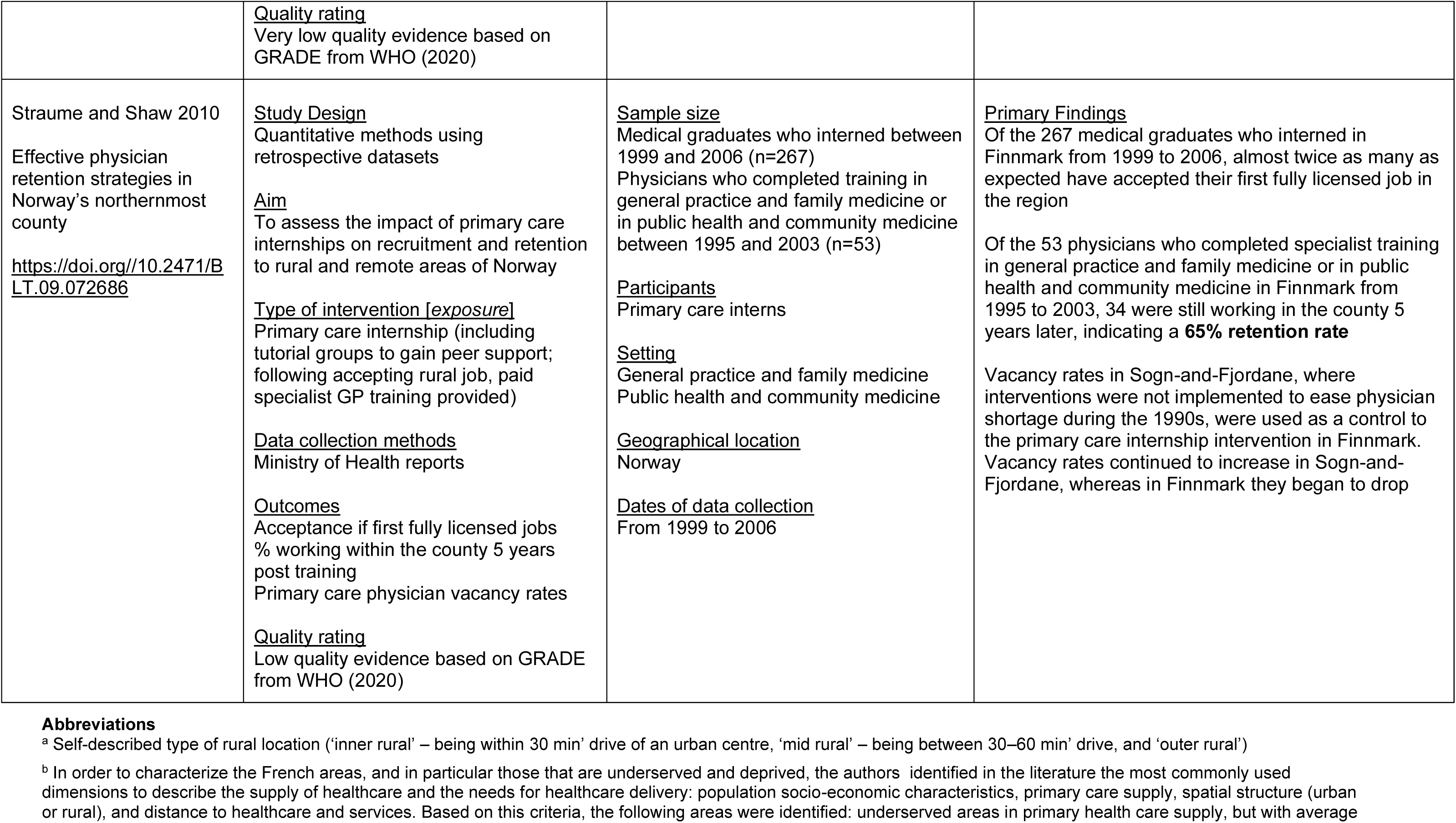
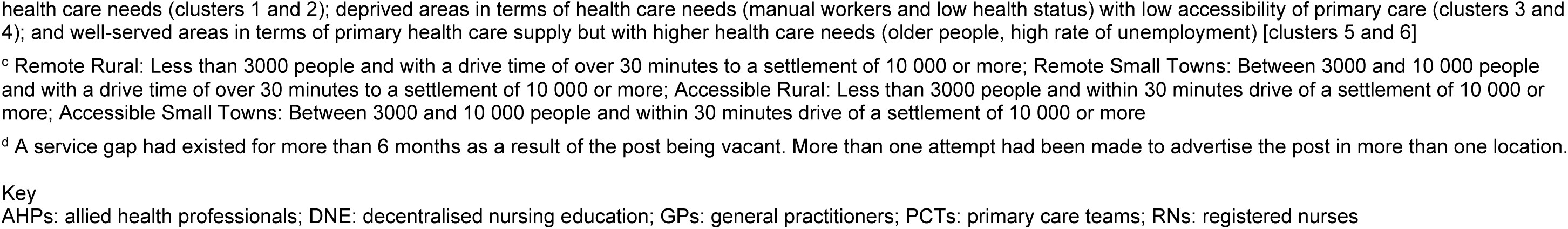
Summary table of primary research studies.

